# Insights from complex trait fine-mapping across diverse populations

**DOI:** 10.1101/2021.09.03.21262975

**Authors:** Masahiro Kanai, Jacob C Ulirsch, Juha Karjalainen, Mitja Kurki, Konrad J Karczewski, Eric Fauman, Qingbo S Wang, Hannah Jacobs, François Aguet, Kristin G Ardlie, Nurlan Kerimov, Kaur Alasoo, Christian Benner, Kazuyoshi Ishigaki, Saori Sakaue, Steven Reilly, The BioBank Japan Project, FinnGen, Yoichiro Kamatani, Koichi Matsuda, Aarno Palotie, Benjamin M Neale, Ryan Tewhey, Pardis C Sabeti, Yukinori Okada, Mark J Daly, Hilary K Finucane

**Author notes:** These authors jointly supervised this work (Mark J Daly, Hilary K Finucane). Corresponding authors: Masahiro Kanai, Mark J Daly, and Hilary K Finucane.

## Abstract

Despite the great success of genome-wide association studies (GWAS) in identifying genetic loci significantly associated with diseases, the vast majority of causal variants underlying disease-associated loci have not been identified^1–3^. To create an atlas of causal variants, we performed and integrated fine-mapping across 148 complex traits in three large-scale biobanks (BioBank Japan^4,5^, FinnGen^6^, and UK Biobank^7,8^; total *n* = 811,261), resulting in 4,518 variant-trait pairs with high posterior probability (> 0.9) of causality. Of these, we found 285 high-confidence variant-trait pairs replicated across multiple populations, and we characterized multiple contributors to the surprising lack of overlap among fine-mapping results from different biobanks. By studying the bottlenecked Finnish and Japanese populations, we identified 21 and 26 putative causal coding variants with extreme allele frequency enrichment (> 10-fold) in these two populations, respectively. Aggregating data across populations enabled identification of 1,492 unique fine-mapped coding variants and 176 genes in which multiple independent coding variants influence the same trait (*i.e.*, with an allelic series of coding variants). Our results demonstrate that fine-mapping in diverse populations enables novel insights into the biology of complex traits by pinpointing high-confidence causal variants for further characterization.

## Introduction

Identifying causal variants for complex traits is a major goal of human genetics research, but most genome-wide association studies (GWAS) do not pinpoint specific variants, limiting the biological inference possible from follow-up experimentation^1–3^. Identifying causal variants from GWAS associations (*i.e.*, fine-mapping) is challenging due to extensive linkage disequilibrium (LD) among associated variants, effect sizes that are often small, and the presence of multiple independent causal variants at a locus. Fine-mapping methods assign to each variant a posterior probability of being a causal variant (posterior inclusion probability, PIP)^9–16^, and recently-developed methods for fine-mapping use scalable, sophisticated algorithms^14–16^ that allow for multiple causal variants in a locus and can be applied to the very large data sets necessary to overcome the challenges listed above. Previous studies, performed almost exclusively in cohorts of European ancestry^17–22^ or meta-analyses of majority European ancestry^23–30^, have used fine-mapping methods to identify putative causal variants, enabling novel biological insights into diseases such as inflammatory bowel disease^19^ and type 2 diabetes^20^ and traits such as blood cell counts^21^ and kidney function^30^.

The recent development of large-scale biobanks worldwide^4,6,7,23^ provides an exciting opportunity for well-powered fine-mapping of multiple phenotypes in diverse populations of both European and non-European ancestries. Unlike results from most meta-analyses, biobanks allow access to individual-level genotypes at large scale, enabling more accurate fine-mapping results^21,22^, and often include hundreds of complex diseases and quantitative traits. For example, BioBank Japan (BBJ)^4,5^, the largest non-European biobank, has recruited ∼200,000 individuals with >200 phenotypes, which is sufficient to achieve powerful fine-mapping in a cohort of East Asian ancestry. Within Europe, there is also substantial genetic diversity^31^; for example, FinnGen^6^, a biobank in Finland, currently combines genotype data with electronic health records for ∼270,000 individuals in a population that has undergone strong population bottleneck followed by subsequent isolation and rapid expansion, making it genetically distinct from mainland Europe^32^. Moreover, because both Japan and Finland have recently undergone population bottlenecks, these populations harbor deleterious alleles with high frequency that are rare or absent in other populations^33–35^.

Here, for the first time, we compare and combine fine-mapping results across large-scale biobanks in three distinct populations. To this end, we apply state-of-the-art multiple-causal-variant fine-mapping methods at scale in BBJ^4,5^ and FinnGen^6^, and we analyze these results in conjunction with results from our parallel effort performing fine-mapping in UK Biobank (UKBB)^7,8^. Our multiple-biobank fine-mapping enables us to identify high-confidence putative causal variants that replicate in multiple populations, to compare fine-mapping results across biobanks, and to identify population-specific putative causal variants and the genes these variants converge on.

## Results

### Expanded atlas of putative causal variants across three populations

In a companion paper^8^, we describe our fine-mapping in UKBB^7^ (*n* = 361,194; 119 traits); here, to create an atlas of causal variants of complex traits, we extended our analysis to additionally include 148 complex diseases and traits available in BBJ^4,5^ (*n* = 178,726; 79 traits) and FinnGen^6^ (*n* = 271,341; 67 traits from release 6) (**Fig. 1a**; **Supplementary Table 1,2**). These traits were manually curated in each biobank to cover a wide spectrum of human phenotypes ranging from common complex diseases to biomarkers. Of these, 26 traits (*e.g.,* height and type 2 diabetes) are available in all the three cohorts, 65 traits (*e.g.*, lab tests and biomarkers) are available in any two of the three, and the rest are specific to a single cohort (**Fig. 1b**). We performed GWAS in BBJ and FinnGen using a generalized linear mixed model as implemented in SAIGE^36^ or BOLT-LMM^37,38^ (**Methods**). We identified 2,611 and 1,698 genome-wide significant locus-trait pairs (*P* < 5.0 × 10^−8^; 3 Mb regions excluding the major histocompatibility complex [MHC]; **Methods**) in BBJ and FinnGen, respectively. We then conducted multiple-causal-variant fine-mapping using FINEMAP^14,15^ and SuSiE^16^ (**Methods**).

**Fig. 1.**
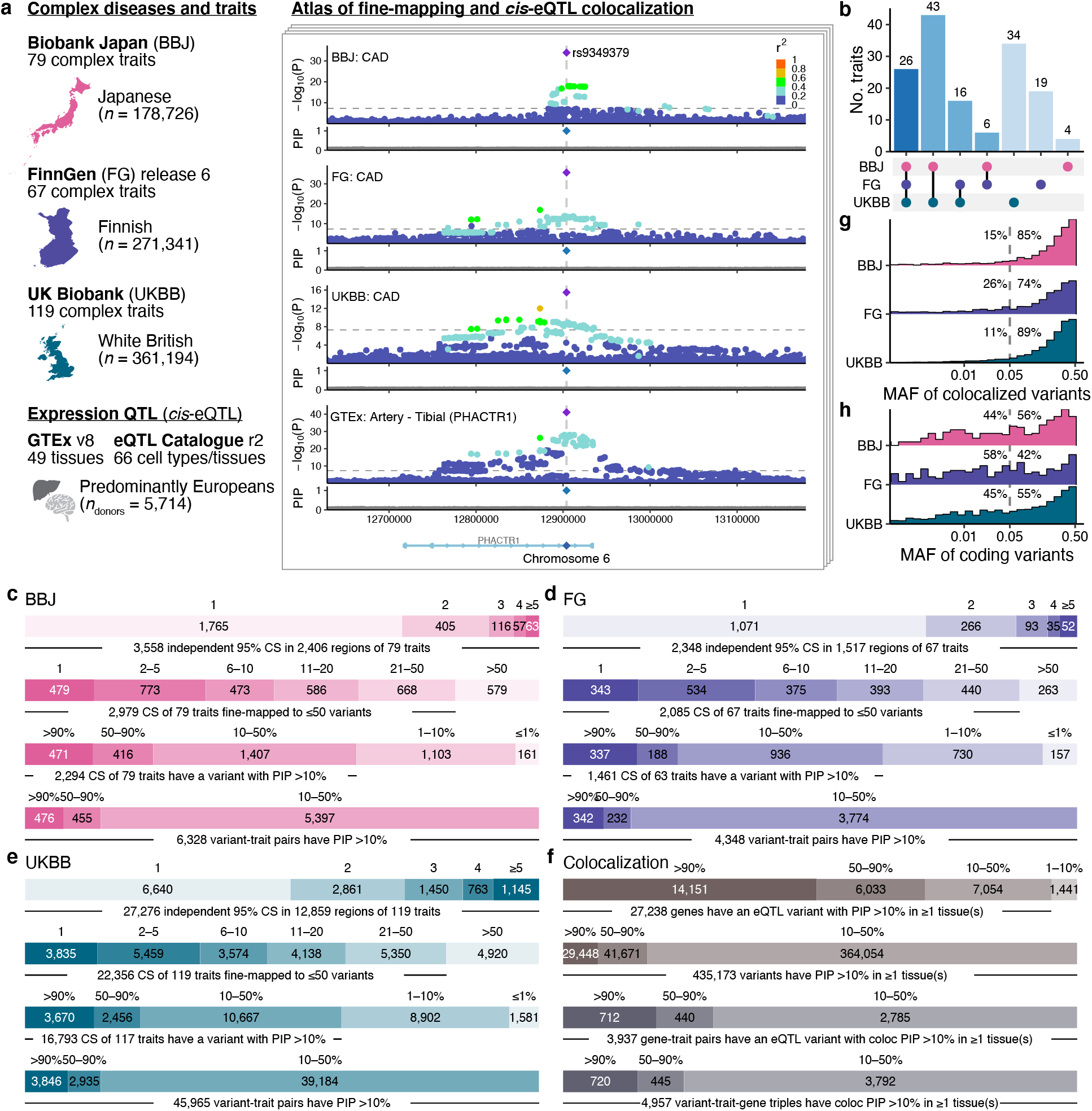
Expanded atlas of putative causal variants across three populations. **a.** Overview of the studied cohorts and *cis*-eQTL datasets. As an illustrative example, the 6p24.1 locus was shown for coronary artery disease (CAD) association in BBJ, FinnGen, and UKBB with *cis*-eQTL association of *PHACTR1* in tibial artery from GTEx. **b.** Number of traits shared across the cohorts. **c–e.** For each cohort, number of independent 95% CS per region, number of fine-mapped variants per 95% CS, number of 95% CS binned by the best PIP variant in each CS, and number of fine-mapped variants binned by PIP. All numbers are counted against unique trait pairs. **f.** (Top two rows) number of genes or variants binned by the best PIP*cis*-eQTL across tissues. (Bottom two rows) number of gene-trait pairs or variant-trait-gene triples binned by the best PIPcoloc across tissues. **g.** MAF distribution of cololocalized variants (the best PIPcoloc > 0.1) in each biobank. **h.** MAF distribution of coding variants (the best PIP > 0.1) in each biobank. Labels represent proportions of variants with MAF > 5% and ≤ 5% in each biobank.

In total, our expanded atlas included 476, 342, and 3,847 fine-mapped variant-trait pairs (posterior inclusion probability [PIP] > 0.9), and 3,558, 2,348, and 27,276 95% credible set (CS)-trait pairs (median CS size = 11, 9, and 12) in BBJ, FinnGen, and UKBB, respectively (**Fig. 1c–e**). These consisted of 4,518 unique variant-trait pairs (PIP > 0.9 in any population) and 31,598 unique 95% CS-trait pairs (median CS size = 12; independent SuSiE CS merged across populations; **Methods**) in aggregate, of which 23,563 CS-trait pairs (75%) contained at least one variant with PIP > 0.1 (**Supplementary Table 3,4**). Notably, our expanded atlas included 66 unique variant-trait pairs (PIP > 0.9 in any population) and 601 CS-trait pairs on the understudied X chromosome. The three biobanks displayed similar and strong enrichment of high-PIP (> 0.9) variants in seven main distinct functional categories (defined as non-overlapping regions; **Methods**): predicted loss-of-function (pLoF), missense, synonymous, 5’/3’ UTR, promoter, and cis-regulatory element (CRE) regions (DNase I hypersensitive sites [DHS] and H3K27ac^39^; **Extended Data Fig. 1a–h; Supplementary Table 5**). In addition, our combined results recapitulated the functional enrichments of 35 additional annotations as previously reported^40–43^, including conserved regions in mammals^44,45^ and ancient putative promoter/enhancer^46^; these enrichments remained significant even when analysis is restricted to the “non-genic” variants that do not belong to any of the seven main functional categories listed above (**Extended Data Fig. 1i; Supplementary Table 6**).

We additionally performed eQTL colocalization in BBJ and FinnGen, using fine-mapped *cis*-eQTLs from GTEx^8,47^ v8 and eQTL catalog^48^ release 4, identifying 719 variant-trait-gene triples; in our companion paper^8^, we identified 4,420 triples in UKBB. We aggregated these results into a combined 4,957 unique variant-trait-gene triples in which the variant was fine-mapped for both the trait and expression of the gene (PIP_coloc_ = PIP_GWAS_ × PIP*_cis_*_-eQTL_ > 0.1), spanning 117 traits and 3,937 genes (**Fig. 1f**; **Supplementary Table 7**). We defined the rate of colocalization as the proportion of variants with PIP > 0.1 in each biobank that showed at least one *cis*-eQTL colocalization (PIP_coloc_ > 0.1 across any trait, gene, or tissue) in our study; this rate was 5.3%, 5.6%, and 7.3% for BBJ, FinnGen, and UKBB, respectively. We investigated the MAF distribution of colocalized variants in each biobank and observed that 85%, 74%, and 89% of colocalized variants showed MAF > 5% in BBJ, FinnGen, and UKBB, respectively (**Fig. 1g**). This is in contrast to the coding variants with PIP > 0.1, of which 56%, 42%, and 55% had MAF > 5% in BBJ, FinnGen, and UKBB, respectively (**Fig. 1h**).

### High-PIP variants are largely non-overlapping across populations

We set out to investigate what proportion of variants with PIP > 0.9 in one population are associated or fine-mapped in other populations. Fine-mapping methods employ a model in which there are a small number of causal variants driving the association signal at the locus, all of which are measured without error, and there are no uncorrected confounding or non-linear effects. When the model is perfectly specified and inference is perfectly accurate, we would expect, for example, 90% of variants with PIP = 0.9 to be truly causal; however, this will not always be the case. We systematically classified variants based on several hierarchical criteria (**Fig. 2a**; **Methods**). First, what proportion of high-PIP (PIP > 0.9) variants in one population (the “discovery population”) reach genome-wide significance (*P*_GWAS_ < 5.0 × 10^−8^) in either of the other two (“secondary”) populations, permitting a well-powered comparison of fine-mapping results at the same locus. Second, of these variants where association is strongly replicated, what proportion have replicated fine-mapping, defined by the same variant having PIP > 0.1 in the secondary population (that is, the variant is also fine-mapped in the second population, though at a lower threshold of confidence). For this analysis, we utilized only the 26 traits analyzed in all three cohorts.

**Fig. 2.**
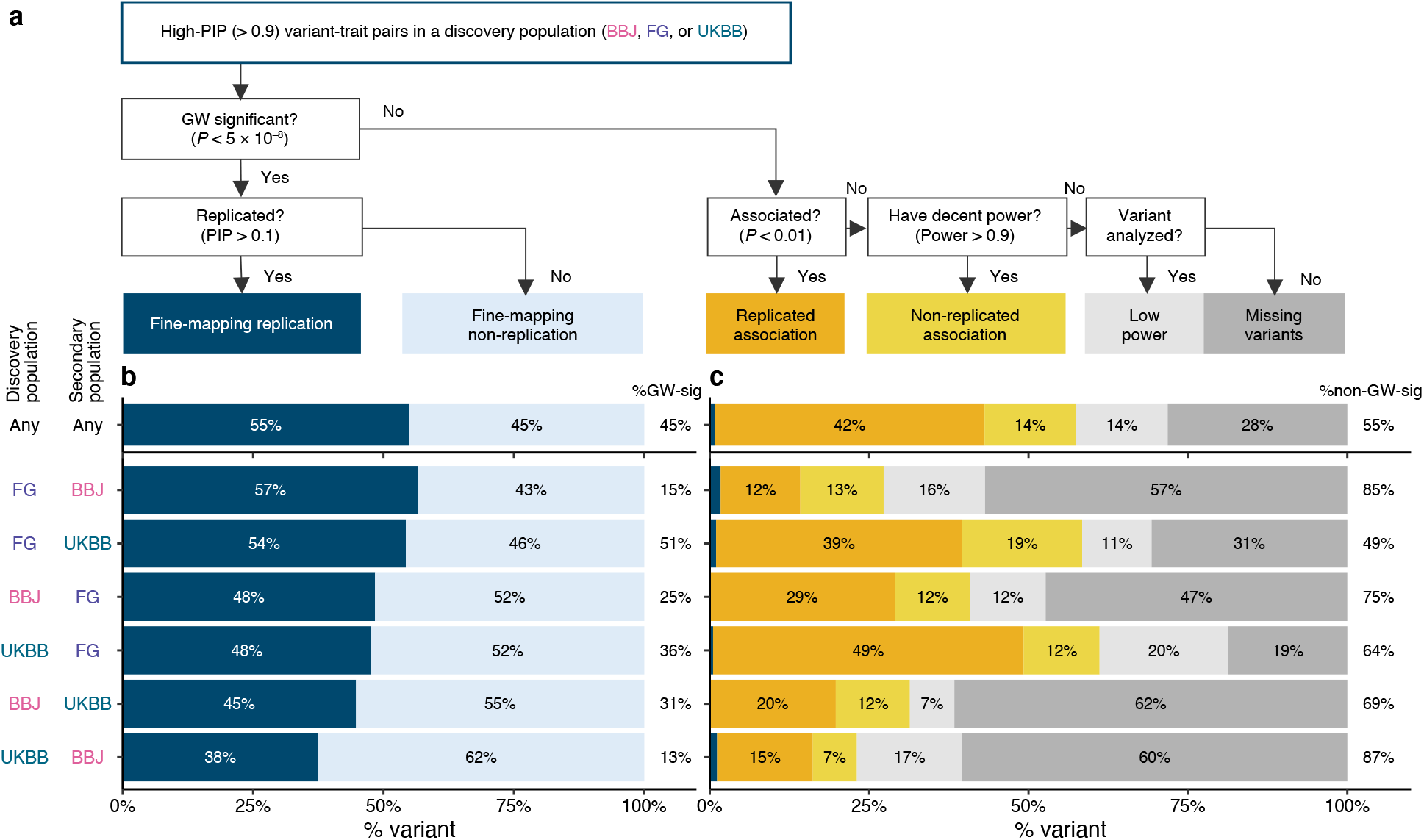
Overview of replication status for high-PIP fine-mapped variants across populations. **a.** Schematic flowchart of our classification criteria. Starting from the high-PIP (> 0.9) variant-trait pairs in a discovery population, we categorized each pair into the six categories: fine-mapping replication, fine-mapping non-replication, replicated association, non-replicated association, low power, and missing variants (**Methods**). **b,c.** Barplots showing a fraction of the high-PIP (> 0.9) variant-trait pairs identified in each discovery population, stratified by the above replication categories tested in the other two secondary populations. Labels in the bar represent a proportion for each category, while labels on the right represent a proportion of the genome-wide significant and non-genome-wide significant variant-trait pairs. **b**. Breakdowns for the genome-wide significant variant-trait pairs (*P*GWAS < 5.0 × 10^−8^) in a secondary population. **c**. Breakdowns for the non genome-wide significant variant-trait pairs (*P*GWAS ≥ 5.0 × 10^−8^) in a secondary population. Note that there were three variant-trait pairs in total that had replicated fine-mapping (PIP > 0.1) but not genome-wide significance in either of the secondary populations (dark blue).

Out of 646 unique variant-trait pairs with PIP > 0.9 in at least one of the three populations, we found that 45% (291 / 646) achieved genome-wide significance (*P*_GWAS_ < 5.0 × 10^−8^) in at least one of the other two populations (**Fig. 2b**). Of these, we found that 55% (160 / 291) had replicating fine-mapping (PIP > 0.1) in at least one of the other two populations, while the other 45% (131 / 291) did not (PIP ≤ 0.1). We took the proportion of fine-mapping replication (= # replication / [# replication + # non-replication] among the variants reaching *P*_GWAS_ < 5.0 × 10^−8^) and defined it as the cross-biobank fine-mapping replication rate. This proportion was relatively consistent across all the pairs of populations, ranging from 38% to 57% (**Fig. 2b**). The cross-biobank fine-mapping replication rate was relatively insensitive to the specific threshold, increasing only slightly when considering a fine-mapping result to be replicated if it had PIP > 0.05 or was in a 95% credible set, as opposed to PIP > 0.1 (**Extended Data Fig. 2a,b**). While mean PIP in a secondary population was positively correlated with PIP in the discovery population, the underlying distribution of PIP in the secondary populations were bimodal, particularly for variants with PIP > 0.9 in the discovery population (**Extended Data Fig. 2c–e**).

To further interpret these observations, we analyzed simulated GWAS data described in our companion paper^8^, and we characterized specific examples. In our simulations, our fine-mapping algorithm was mostly well calibrated, with 96% of variants with PIP>0.9 truly causal. In these simulations, however, the variants that were simulated to be causal and that reached genome-wide significance also had a bimodal distribution of PIP, with 24% reaching PIP>0.9, due to incomplete power for fine-mapping. Thus, while we have the highest confidence in the fine-mapping results that replicate across populations, we do not interpret a cross-biobank replication rate of 55% as strong evidence for or against fine-mapping miscalibration. In examining specific examples in real data, we found that lack of replication was sometimes due to differences in LD structure and effect sizes across populations that lower power in the secondary population, or likely non-causal variants that nonetheless achieve high PIP in the discovery population, as expected given the PIP threshold of 0.9. We illustrated a few examples in **Extended Data Fig. 3**.

Of the remaining 55% (355 / 646) of variant-trait pairs that did not reach genome-wide significance (*P*_GWAS_ < 5.0 × 10^−8^) in either of the secondary populations, 42% (150 / 355) had an association that replicated at the more permissive threshold of *P*_GWAS_ < 0.01 (**Fig. 2c**), suggesting the association is present but at a level insufficient to perform fine-mapping reliably. An additional 14% (51 / 352) had high power to detect association (power > 0.9 for achieving *P*_GWAS_ < 0.01; **Methods**) in at least one of the secondary populations, assuming the same causal effect size from the discovery population and a standard linear regression, but were not associated at *P*_GWAS_ < 0.01 in either population. These variants may include causal variants with heterogeneous effect sizes across populations or false positive variants that nonetheless achieved high PIP in the discovery population (which is not unexpected given the number of traits studied and the PIP threshold of 0.9). A few causal variants would also be expected not to reach this threshold due only to random sampling, even with equal effect sizes and estimated power of 0.9. We note that three variant-trait pairs had replicated fine-mapping (PIP > 0.1) but not genome-wide significance in either of the secondary populations (Note that these are in genome-wide significant loci). Lastly, 42% (151 / 355) had low power or were missing from the GWAS summary statistics due mostly to differences in allele frequencies across populations (**Fig. 2c**; **Supplementary Note**). This proportion was different for different pairs of populations, ranging from 19% (UKBB and FinnGen) to 62% (BBJ and UKBB). Importantly, our results indicate that these missing causal variants are *undiscoverable* through standard GWAS fine-mapping in other populations, re-emphasizing the desperate need for data generation in diverse populations.

For the remainder of this manuscript, we mainly focus on several subsets of PIP > 0.9 variants with highest confidence: fine-mapped variants replicated in multiple populations, coding variants with PIP > 0.9, and genes supported by multiple fine-mapped variants.

### Common putative causal variants implicate shared biological mechanisms across populations

Restricting to 91 traits available in two or more populations, we identified 285 high-confidence variant-trait pairs (204 unique variants including 56 variants that are only polymorphic in Europeans) that achieve replicated fine-mapping across multiple populations analyzed (PIP > 0.9 in at least one population and PIP > 0.1 in at least one of the others; **Supplementary Table 8,9**). While estimating correlation of effect sizes across multiple traits and populations is challenging, we observed 100% directional consistency for posterior effect sizes between populations (*P* for sign test = 2.7 × 10^−107^). These replicated fine-mapped variants represent a set of common putative causal variants (**Extended Data Fig. 4a,b**) with the highest confidence in our dataset, providing excellent candidates for functional characterization and therapeutic targets.

We observed a significant enrichment of coding variants in high-confidence variant-trait pairs: of the 285 high-confidence variant-trait pairs, 94 pairs (60 unique variants) are coding variants (**Supplementary Table 8**), whereas 4 pairs would be expected by chance (Fisher’s exact test *P* < 0.05). These variants include well-known pLoF and missense variants such as rs429358 (*APOE* ε4-tagging missense variant) for Alzheimer’s disease^49^; rs2066847 (*NOD2*: p.Leu980ProfsTer2) for Crohn’s disease^50,51^; rs855791 (*TMPRSS6*: p.Val736Ala) for blood hemoglobin levels and erythrocyte volume^52^; rs2642438 (*MARC1*: p.Ala165Thr) for alkaline phosphatase^53,54^; and rs4149056 (*SLCO1B1*: p.Val174Ala) for total bilirubin^55^. Notably, we found that rs9379084 (*RREB1*: p.Asp1171Asn) showed PIP > 0.9 for height in every population; this variant was previously implicated for type 2 diabetes^20^ but not for height. We also found that a common synonymous variant rs55714927 on *ASGR1* (canonical transcript ENST00000269299.3) was fine-mapped for alkaline phosphatase in both BBJ and UKBB (PIP = 1.0 for both; **Extended Data Fig. 5a**). The same variant was significantly associated with other traits in our dataset, such as albumin, cholesterol levels, and sex hormone binding globulin (**Extended Data Fig. 5b**). *ASGR1* was previously reported for having a rare non-coding 12-base-pair deletion within intron 4 (del12; c.284-36_283+33delCTGGGGCTGGGG, NM_001671.4; MAF = 0.41% in ∼398,000 Icelanders), which was associated with a reduced risk of coronary artery disease (CAD), lowering LDL cholesterol, and increasing alkaline phosphatase and vitamin B_12_ levels^56^. However, the reported del12-tagging variant rs186021206 is independent from the synonymous variant rs55714927 (*r*^2^ = 0.001 in Europeans) and is monomorphic in East Asians, implying that the del12 variant does not contribute to the identified rs55714927 association here. Instead, we observed rs55714927 has a significant splicing QTL effect in GTEx liver^47^ (*P* = 2.4 × 10^−46^) for the same isoform as del12 (**Extended Data Fig. 5c,d**).

We also characterized 191 non-coding variant-trait pairs (144 unique variants) with replicated fine-mapping as described above (**Supplementary Table 9**). These variants are primarily located within CRE (48%) followed by promoter (16%) and 3’ UTR (8%) regions, and are enriched for predicted *cis*-regulatory expression modifier score^57^, suggesting that most of these variants act through transcriptional or by post-transcriptional regulation (**Extended Data Fig. 4b–d**). In total, we identified 48 out of 144 putative causal non-coding variants that co-localized with *cis*-eQTL associations (PIP_coloc_ > 0.1 in at least one tissue; **Supplementary Table 9**), including well-known variants, *e.g.*, rs2070895 (intronic variant of *LIPC*) for HDL cholesterol; and rs78378222 (3’ UTR variant of *TP53*) for skin cancer; as well as under-characterized variants, *e.g.*, rs1497406 (intergenic variant, ∼22 kb upstream of *EPHA2*) for γ-glutamyl transferase; and rs34778241 (intronic variant of *EIF4E3*) for loss of Y chromosome (**Extended Data Fig. 6**). Notably, we identified a well-known intronic variant rs9349379 in *PHACTR1* that was fine-mapped for CAD in every population (**Fig. 1b**; PIP = 1.0; MAF = 0.35, 0.45, and 0.41 for BBJ, FinnGen, and UKBB, respectively). This intronic variant also co-localized with a fine-mapped *cis*-eQTL association of *PHACTR1* in GTEx tibial artery^47^ (PIP_coloc_ = 1.0). We note that, although we identified rs9349379 as a putative causal non-coding variant for CAD with high confidence, it was previously demonstrated that rs9349379 also regulates expression of *EDN1* (located a 600 kb upstream of *PHACTR1*) in CRISPR-edited endothelial cells^58^; and thus the causal gene(s) for CAD at this locus remains unresolved^58,59^.

The 144 putative causal non-coding variants also included seven intergenic variants located in gene deserts; *i.e.*, that are more than 250 kb away from the closest gene^60^ (**Supplementary Table 10**). For example, rs77541621 and rs183373024 (349kb and 322kb upstream of *POU5F1B*, respectively) were fine-mapped for prostate cancer (PIP = 1.0 in FinnGen and UKBB), and are located within the 8q24 locus, a well-known gene desert associated with many complex diseases^61,62^ (**Extended Data Fig. 7a**). These variants are one of the 12 independent variants for prostate cancer that were previously identified at the 8q24 locus, but the exact functional mechanism of each variant is still under active investigation^63^. Other examples include rs1434282 (284 kb downstream of *PTPRC*) for mean corpuscular volume, rs116376456 (269 kb downstream of *IRS1*) for height, and rs35009121 (1.2 Mb downstream of *GATA3*) for serum calcium levels (**Extended Data Fig. 7b–d**). Although these loci are also known as gene deserts, none of the fine-mapped variants are well-characterized in the current literature, nor do they overlap with enhancer-gene mappings predicted by the activity-by-contact (ABC) model^64^.

We also found nine examples where a variant was fine-mapped in every population even though it was not significantly associated in every population. Five of these were significant at a more permissive threshold of *P* < 1.0 × 10^−5^, but in other cases the marginal effect sizes were substantially lower, due to LD with another causal variant(s). For example, rs244711 (4.7 kb upstream of *FGFR4*) is consistently fine-mapped for height but not significantly associated in BBJ (marginal *β* = 9.0 × 10^−3^; *P* = 4.1 × 10^−4^; **Extended Data Fig. 8a–d**). We found that rs244711 is partially correlated with a nearby fine-mapped missense variant rs1966265 (*FGFR4*: p.Val10Ile) in every population (*r*^2^ = 0.14, 0.08, and 0.13 in BBJ, FinnGen, and UKBB, respectively) but the correlation is only negative in BBJ (*r* = –0.37). The causal effect of rs244711 is thus partially cancelled out by the tagged effect of rs1966265 in BBJ, where the correlation between the two variants is negative, but not in UKBB and FinnGen, where the correlation is positive, leading to a non-significant association in BBJ but significant associations in UKBB and FinnGen. Another example is rs1801706 (3’ UTR variant of *CETP*), which is consistently fine-mapped for HDL cholesterol but not significantly associated in BBJ (marginal *β* = 6.0 × 10^−3^; *P* = 0.43; **Extended Data Fig. 8e–h**). This is owing to partial correlation with Japanese-enriched splice donor and missense variants rs5742907 (c.1321+1G>A) and rs2303790 (p.Asp459Gly). These two variants showed large effect sizes (marginal *β* = 0.76 and 0.39; *P* = 4.9 × 10^−122^ and 5.5 × 10^−206^; respectively) and are negatively correlated with rs1801706 in BBJ (*r* = –0.03 and –0.06, respectively; this corresponds to –16.6 and –56.3 decrease in marginal *χ*^2^ statistics of rs1801706 by partial tagging). These examples illustrate that, when a region contains multiple independent associations, differences in LD between two sites can create differences in the marginal effect size and observed association in univariate analyses between populations.

### Identification of population-enriched putative causal variants

Given that a substantial number of the variants with high PIP (> 0.9) in one population are rare/absent (and therefore undiscoverable) in the other populations (**Fig. 2c**), we investigated allele frequency (AF)-enriched variants from the two bottlenecked populations included in our study, Finland^65,66^ and Japan^67,68^. To quantify AF enrichment (AFE) in the Finnish and Japanese populations, we used the gnomAD^69^ v2 and the GEM-J WGS^70^ to compute a ratio of AF in Japanese vs. non-Japanese-Korean East Asians (NJKEA) for BBJ and in Finnish vs. non-Finnish-Swedish-Estonian Europeans (NFSEE) for FinnGen (**Methods**).

Past studies have noted that variants stochastically boosted through a bottleneck are enriched for functional categories^33–35,71–73^. Consistent with these previous studies, we found that there were significantly more variants with AFE > 10 than with AFE < 1/10 in both FinnGen and BBJ, and that variants with AFE > 10 were enriched for coding variants (2.2- and 4.8-fold enrichment over variants with AFE ≤ 10; **Methods**). Of 140,416 and 91,564 coding variants tested in FinnGen and BBJ GWAS, 29,656 (21%) and 14,802 (16%) showed AFE > 10 in the Finnish or Japanese population, respectively (**Fig. 3a,b**). Furthermore, high-PIP (> 0.9) coding variants were significantly more likely to have high AFE than low-PIP (≤ 0.01) coding variants (**Fig. 3c,d**; Fisher’s exact test *P* < 0.05; **Methods**); and showed substantially younger estimated allele age based on GEVA^74^ (**Fig. 3e,f**). These observations are consistent with recent bottleneck events and negative selection on the putative causal variants studied here, because deleterious variants boosted in frequency through these bottlenecks have had insufficient time to be brought back down in frequency by selection^75,76^.

**Fig. 3.**
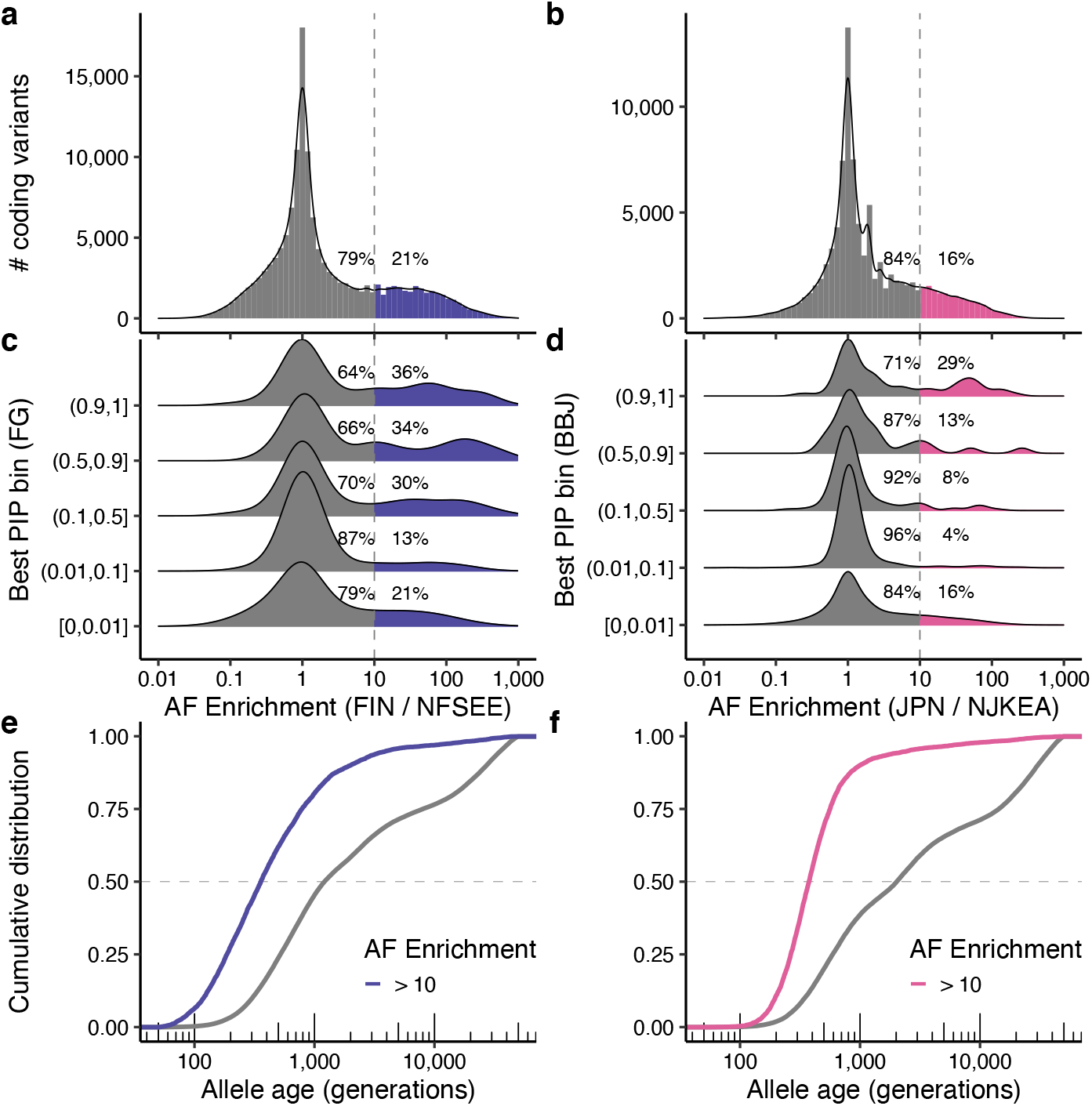
Population-enriched putative causal coding variants. **a–d.** Histograms showing a distribution of allele frequency (AF) enrichment metric in (**a**) Finnish (*n* = 10,824) and (**b**) Japanese (*n* = 7,609) populations. A ratio of AF was computed against NFSEE (*n* = 43,697) and NJKEA (*n* = 7,212) for coding variants analyzed in BBJ or FinnGen GWAS that exist in gnomAD WES or GEM-J WGS. For a subset of variants that are fine-mapped in our analysis (see **Methods**), we show AF enrichment distribution across maximum PIP bins computed in (**c**) FinnGen or (**d**) BBJ. **e–f**. Cumulative distribution of estimated allele age for coding variants, stratified by AF enrichment in (**e**) Finnish or (**f**) Japanese. FIN: Finnish, JPN: Japanese, NFSEE: Non-Finnish-Swedish-Estonian European, NJKEA: Non-Japanese-Korean East Asian.

Notably, we identified seven pLoF variants and 40 missense high-PIP (> 0.9) variants with extreme AF enrichment (> 10-fold) in BBJ or FinnGen (**Table 1**). These variants are more likely to be deleterious and impactful given their extreme enrichment. Indeed, the list includes several known pathogenic variants or genes in related autosomal recessive disorders. For example, rs75326924, a Japanese-enriched missense variant (p.Pro90Ser) on *CD36* is a known pathogenic variant for platelet glycoprotein IV (CD36) deficiency (PIP = 1.0 for platelet count; MAF = 0.047 in GEM-J WGS), contributing to high prevalence of CD36 deficiency in Japanese (2– 3%)^77^; and rs386833873, a Finnish-enriched frameshift variant (p.Leu41AspfsTer50) on *NPHS1* is a well-known causal variant for the congenital nephrotic syndrome of the Finnish type (PIP = 1.0 for nephrotic syndrome; MAF = 0.011 in gnomAD Finnish)^78^. Interestingly, we found two novel population-enriched deleterious variants on *PLOD2*, fine-mapped for height: i) a Japanese-enriched missense variant rs148051196 (p.Gln553Arg; PIP = 1.0; MAF = 7.3 × 10^−3^ in GEM-J WGS) and ii) a Finnish-specific stop-gained variant rs201501322 (p.Ser166Ter; PIP = 0.58; MAF = 1.9 × 10^−3^ in gnomAD FIN). *PLOD2* is a known recessive gene for Bruck syndrome 2 (osteogenesis imperfecta with congenital joint contractures; OMIM: 609220)^79^. We identified additional population-enriched variants for height in 27 genes, including known recessive genes such as *ADAMTS17* (causal gene for Weill-Marchesani syndrome 4; OMIM: 613195) and *IHH* (brachydactyly type A1; OMIM: 112500). Furthermore, we identified fine-mapped variants on genes that were not previously implicated, such as rs199935580 (*THBS3*: p.Arg520Trp; MAF = 1.0 × 10^−3^ in gnomAD FIN) fine-mapped for carpal tunnel syndrome (PIP = 1.0); rs191692991 (*LUM*: p.Arg310Cys; MAF = 5.5 × 10^−3^ in gnomAD FIN) fine-mapped for fibroblastic disorders (PIP = 1.0); and rs200939713 (*POF1B*: p.Arg339Trp; MAF = 1.7 × 10^−3^ in gnomAD FIN) fine-mapped for varicose veins (PIP = 0.99). Detailed biological annotations of each gene are summarized in the **Supplementary Box**.

**Table 1.** Population-enriched putative causal coding variants. Nonsynonymous coding variants (PIP > 0.9) with AFE > 10 in the Japanese or Finnish populations are shown.

| Variant | rsid | Gene | Consequence | AF (pop) | AF (ref) | AFE | Best PIP | Fine-mapped traits (PIP > 0.9) |
| --- | --- | --- | --- | --- | --- | --- | --- | --- |
| **BBJ** |  |  |  |  |  |  |  |  |
| 1:21890590:G:A | rs199669988 | ALPL | Missense | 0.015 | 0.00035 | 43.1 | 1 | ALP |
| 1:55505604:G:A | rs564427867 | PCSK9 | Missense | 0.012 | NA | Inf | 1 | LDLC, TC |
| 2:21242731:G:A | rs13306206 | APOB | Missense | 0.039 | 0.00049 | 79.5 | 1 | LDLC, MI, TC |
| 2:44051573:T:TA | rs142037828 | ABCG5 | Splice region | 0.051 | 0.00042 | 123.2 | 1 | Cholelithiasis |
| 2:120231070:C:G | rs3731600 | SCTR | Missense | 0.048 | 0.00069 | 69.2 | 0.99 | T2D |
| 2:219919943:C:T | rs200216644 | IHH | Missense | 0.0036 | 0.00027 | 13.4 | 0.99 | Height |
| 3:145794588:T:C | rs148051196 | PLOD2 | Missense | 0.0073 | 0.00014 | 52.4 | 1 | Height |
| 4:6303731:G:A | rs147834269 | WFS1 | Missense | 0.053 | 0.0012 | 43.7 | 1 | T2D |
| 6:158484904:G:C | rs141160611 | SYNJ2 | Missense | 0.032 | 0.0025 | 12.9 | 0.96 | GGT |
| 7:45954540:C:T | rs17847676 | IGFBP3 | Missense | 0.0061 | 6.90E-05 | 88.5 | 1 | Height |
| 7:80286003:C:T | rs75326924 | CD36 | Missense | 0.047 | 0.00083 | 56.9 | 1 | Plt |
| 8:118184855:A:T | rs770224130 | SLC30A8 | Missense | 0.0062 | 0.00014 | 44.7 | 0.97 | T2D |
| 9:107593923:C:G | rs754040394 | ABCA1 | Missense | 0.0019 | NA | Inf | 1 | HDLC, TC |
| 11:64361219:G:A | rs121907892 | SLC22A12 | Stop gained | 0.021 | 0.00042 | 51.6 | 1 | UA |
| 11:116661394:G:C | rs201229911 | APOA5 | Missense | 0.01 | NA | Inf | 1 | HDLC, TG |
| 12:16510581:GAA:G | rs779999476 | MGST1 | Frameshift | 0.015 | NA | Inf | 1 | HDLC |
| 12:109690842:C:T | rs17848833 | ACACB | Missense | 0.0032 | NA | Inf | 1 | HDLC, TG |
| 16:57016150:G:A | rs5742907 | CETP | Splice donor | 0.0037 | NA | Inf | 1 | HDLC, TC |
| 16:84872195:G:C | rs965984074 | CRISPLD2 | Missense | 0.00086 | NA | Inf | 0.99 | Height |
| 17:7462468:G:T | rs201860460 | TNFSF13 | Missense | 0.0022 | 7.00E-05 | 31.1 | 1 | AG, NAP |
| 17:48545926:C:A | rs201158957 | CHAD | Missense | 0.0081 | 6.90E-05 | 116.1 | 1 | Height |
| 17:78358945:G:A | rs112735431 | RNF213 | Missense | 0.01 | 0.00049 | 21.2 | 1 | CAD, MAP, PP, SBP |
| 19:11241988:C:T | rs13306505 | LDLR | Missense | 0.0085 | 0.00021 | 41.1 | 1 | LDLC, TC |
| 19:42855705:G:A | rs200485103 | MEGF8 | Missense | 0.0039 | NA | Inf | 0.95 | Glucose |
| 19:46178043:G:T | rs13306398 | GIPR | Missense | 0.02 | 6.90E-05 | 286.9 | 1 | BMI, BW |
| 20:44507112:G:A | rs139396693 | ZSWIM3 | Missense | 0.015 | 0.00014 | 106.6 | 1 | MCV |
| **FG** |  |  |  |  |  |  |  |  |
| 1:21890632:G:A | rs121918007 | ALPL | Missense | 0.017 | 0.0011 | 15.7 | 0.94 | Urolithiasis |
| 1:155170392:G:A | rs199935580 | THBS3 | Missense | 0.001 | 1.10E-05 | 88.9 | 1 | Carpal_Tunnel_Syndrome |
| 1:192779303:G:T | rs201233692 | RGS2 | Missense | 0.0088 | 1.10E-05 | 761.5 | 0.96 | Statin |
| 4:120528397:C:T | rs202226125 | PDE5A | Missense | 0.007 | 1.20E-05 | 612 | 1 | Height |
| 5:1272362:G:A | rs770066110 | TERT | Stop gained | 0.00052 | NA | Inf | 1 | IPF |
| 5:1279485:T:C | rs776981958 | TERT | Missense | 0.0016 | NA | Inf | 0.96 | IPF |
| 6:155450779:A:G | rs148543891 | TIAM2 | Missense | 0.031 | 6.90E-05 | 455.3 | 1 | Height |
| 9:35609378:C:T | rs777777413 | TESK1 | Missense | 0.0025 | 2.40E-05 | 101.4 | 1 | Height |
| 9:136501728:C:T | rs77273740 | DBH | Missense | 0.051 | 0.0023 | 21.7 | 1 | Hypertension |
| 10:13040400:A:G | rs199848893 | CCDC3 | Missense | 0.0021 | NA | Inf | 1 | Height |
| 11:36248678:T:TG | rs767680853 | LDLRAD3 | Frameshift | 0.0019 | 2.30E-05 | 82.3 | 1 | Height |
| 12:6882498:C:A | rs149722682 | LAG3 | Missense | 0.00061 | NA | Inf | 1 | AID, Hypothyroidism |
| 12:91498031:G:A | rs191692991 | LUM | Missense | 0.0053 | 1.20E-05 | 450.7 | 1 | Fibroblastic_Disorders, Height |
| 14:100134609:G:A | rs201483470 | HHIPL1 | Missense | 0.0093 | 0.00013 | 74.2 | 0.97 | Height |
| 15:28228553:C:T | rs74653330 | OCA2 | Missense | 0.048 | 0.0014 | 33.4 | 1 | Malignant_Neoplasms, SkC |
| 15:101569374:C:T | rs41531245 | LRRK1 | Missense | 0.0076 | 0.00073 | 10.4 | 1 | Fibroblastic_Disorders, Inguinal_Hernia |
| 17:56436130:C:T | rs199598395 | RNF43 | Missense | 0.012 | 5.00E-05 | 239.7 | 1 | Iron_Deficiency_Anaemia |
| 17:60493445:C:T | rs552441218 | EFCAB3 | Stop gained | 0.001 | 6.90E-05 | 14.7 | 0.98 | Depression_medications |
| 19:36342510:CAG:C | rs386833873 | NPHS1 | Frameshift | 0.011 | 2.40E-05 | 473.3 | 1 | Nephrotic_Syndrome |
| 19:58421417:ACT:A | rs774674736 | ZNF417 | Frameshift | 0.0018 | 4.60E-05 | 39.8 | 0.93 | Chronic_Tonsillitis |
| X:84563165:G:A | rs200939713 | POF1B | Missense | 0.0016 | NA | Inf | 0.99 | Varicose_Veins |

On the other hand, the high-PIP non-coding variants were not significantly more likely to have high AFE than low-PIP non-coding variants (**Extended Data Fig. 9**; Fisher’s exact test *P* > 0.05),partly because non-coding variants tend to be less deleterious and thus less likely to undergo strong negative selection. However, we identified 23 population-enriched (> 10-fold) high-PIP (> 0.9) non-coding variants that are independent of population-enriched coding variants (*r*^2^ < 0.1) in each population (**Supplementary Table 11**). While we are not able to replicate these population-enriched variants in other populations due to low AF, we identified several variants that might have biological significance. For example, a Finnish-enriched rs748670681 in an intron of *TNRC18* (MAF = 0.042 in gnomAD FIN) is fine-mapped for inflammatory bowel disease (IBD) and psoriasis (PIP = 1.0). Despite very significant association in FinnGen (*P* = 6.2 × 10^−69^ for IBD), this locus was not previously reported, and its biological function is not well-characterized.

### Allelic series of putative causal variants across populations

Given that many fine-mapped variants are population-specific, we aggregated results across populations to identify genes harboring fine-mapped coding variants for one or more traits. Overall, we identified 1,492 unique putative causal pLoF/missense variants (best PIP > 0.1) that mapped onto 1,113 genes (**Supplementary Table 12**). Of these genes, 240 have two or more putative causal pLoF/missense variants located on the same gene, and 113 have variants identified from multiple populations (**Fig. 4a**). The genes with the most putative causal pLoF/missense variants include *APOB* (13 missense variants; the loss-of-function observed/expected upper bound fraction [LOEUF]^69^ = 0.46), *TFR2* (1 pLoF and 6 missense variants; LOEUF = 0.77), and *PIEZO1* (7 missense variants; LOEUF = 0.58); despite containing many variants that impact human phenotypes, these genes are modestly constrained (**Fig. 4b, Extended Data Fig. 10a,b**).

**Fig. 4.**
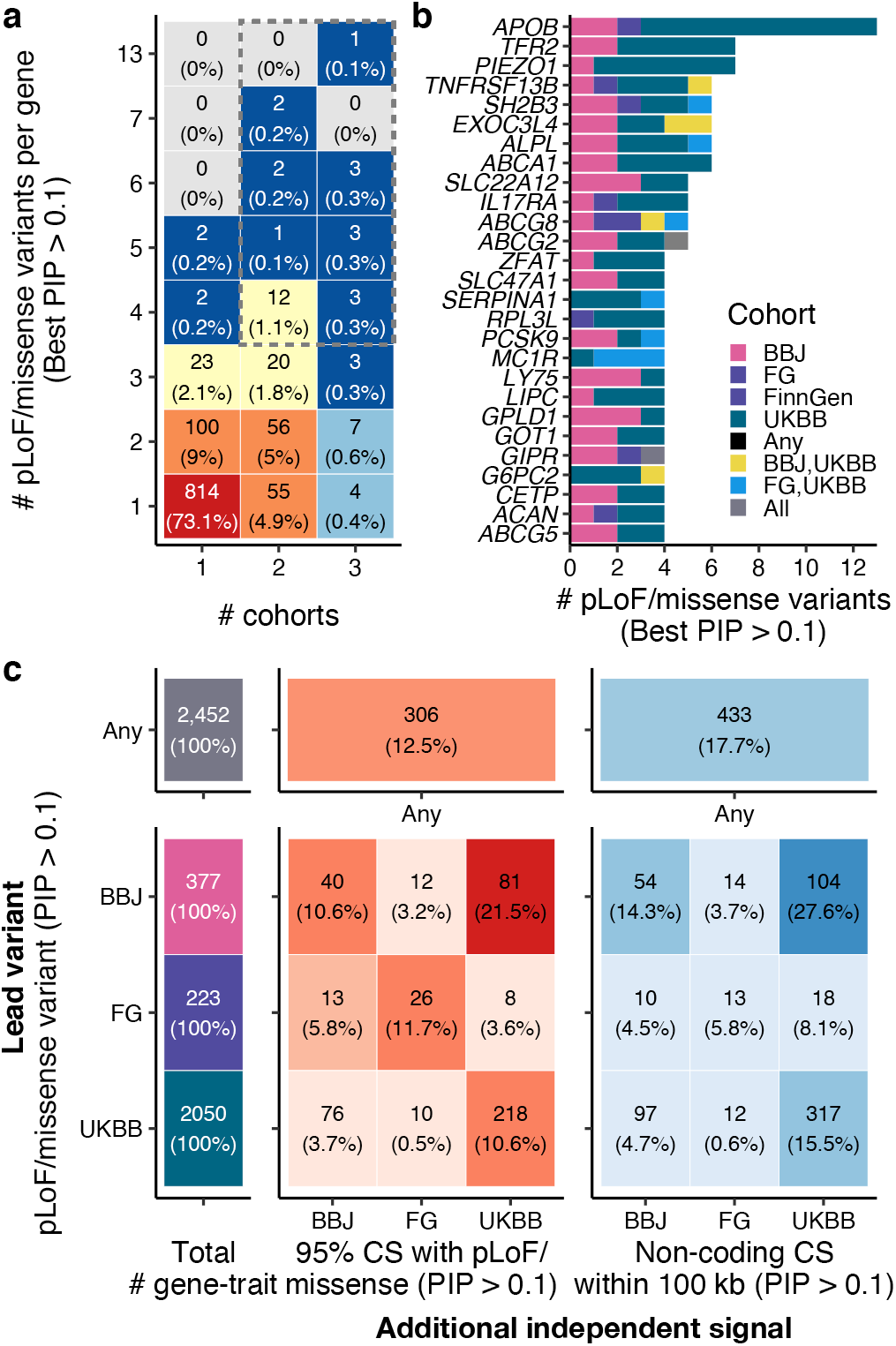
Allelic series of putative causal variants across multiple populations. **a.** Number of fine-mapped pLoF/missense variants (best PIP > 0.1) per gene, stratified by the number of cohorts identified. **b.** Top list of genes that have a large number of fine-mapped pLoF/missense variants (best PIP > 0.1). **c.** Number of additional independent signals identified for a gene with fine-mapped pLoF/missense variants (PIP > 0.1), stratified by a discovery cohort. For each gene-trait pair where we fine-mapped pLoF/missense variants, we counted how many additional independent 95% CS with pLoF/missense and non-coding variants (PIP > 0.1) were identified for the same gene in each cohort.

Next, we focused on allelic series in which multiple fine-mapped coding variants implicate the same gene-trait pair (**Fig. 4c**). There were 2,452 gene-trait pairs with at least one fine-mapped pLoF/missense variant (PIP > 0.1), of which 306 pairs (176 unique genes) had two or more independent pLoF/missense variants (PIP > 0.1), forming an allelic series (**Supplementary Table 13**). We found 104 allelic series (69 unique genes) that included variants fine-mapped in multiple populations, of which 41 allelic series (34 unique genes) included at most one variant per population, making them discoverable only by aggregating data across populations. The cross-population allelic series include e.g., *ABCG2*, a known pathogenic gene for gout, where we identified two pLoF/missense variants (p.Gln126Ter and p.Phe489Leu) in BBJ, two missense variants (p.Asp620Asn and p.Ala528Thr) in UKBB, and one missense variant (p.Gln141Lys) in BBJ, FinnGen, and UKBB (**Extended Data Fig. 10c**).

We further investigated allelic series including both coding and non-coding variants, assuming that non-coding causal variants proximal to deleterious coding variants (< 100 kb) might act through regulation of the same gene^80^. This facilitates understanding of unknown non-coding functions and enables us to identify allelic series for an additional 263 gene-trait pairs (195 unique genes) through coding/non-coding allelic series, of which 107 pairs (87 unique genes) included variants fine-mapped across multiple populations (**Supplementary Table 14**). For example, we identified coding/non-coding allelic series around *EPX* (eosinophil peroxidase) for eosinophil count (**Extended Data Fig. 10d**), where we found European-specific missense variant rs149610649 (*EPX*: p.Phe308Leu; MAF = 0.083 in gnomAD NFE) and Japanese-specific intergenic variant rs536070968 (MAF = 0.011 in GEM-J WGS). The intergenic variant rs536070968 is located 33 kb downstream of *EPX* and 11 kb upstream of *LPO* (lactoperoxidase), an ortholog of *EPX*, illustrating the value of allelic series across multiple populations to assign a potential causal gene from nearby genes.

## Discussion

In this study, we performed statistical fine-mapping in Biobank Japan and FinnGen, and aggregated these results with our parallel fine-mapping of UK Biobank^8^, providing an extensive list of candidate causal variants for 148 complex diseases and traits across diverse populations. By integrating fine-mapped variants from deeply-phenotyped biobanks and eQTL studies, we expanded both the depth and breadth of the resource to explore biological mechanisms of complex traits at single-variant resolution, with replication across multiple populations and colocalization with different tissues. We make these resources publicly available for the community to further accelerate variant prioritization and characterization.

Examination of fine-mapping from three biobanks enabled the identification of 285 high-confidence variant-trait pairs that are replicated across multiple populations. However, the majority of high-PIP (> 0.9) variants are non-overlapping across populations. Many of the variants with high PIP in one population but not in the other two populations were trivially explained by the fact they are rare or monomorphic in the other two populations. The abundance of population-enriched variants exemplifies the significant value of diverse populations in fine-mapping studies, contributing to identification of population-specific discoveries and deeper allelic series of multiple variants at the same locus across populations. We also classified observed inconsistencies into those with potential heterogeneity in effect sizes (*i.e.*, population-specific effects) and those with replicated GWAS association but without replicated fine-mapping, further guiding interpretation of these results.

Our study has several limitations that suggest directions for future work. First, the current fine-mapping methods rely on modeling assumptions that are not all met in real-world fine-mapping (*e.g.*, no genotyping or imputation errors). While we have focused here on a high confidence subset of results—high-PIP variants that replicate across biobanks, and fine-mapped coding variants—we see further exploration of potential misspecification of fine-mapping models as an important area for future work. Second, our sample sizes are still limited, especially for non-European populations, emphasizing the desperate need for more diversity in human genetics. Here, we were powered to fine-map variants with large or moderate effect sizes; more samples will be required to fine-map causal variants with small effect sizes. Moreover, molecular data from non-European samples are vastly limited, which fundamentally inhibits variant interpretation of population-enriched variants. Third, systematic differences in study design, genotyping and imputation across cohorts limited our ability to integrate data from the three biobanks. We see method development for cross-population fine-mapping that takes into account this heterogeneity as an important direction for future work.

We note that the populations studied here differ not only by ancestry, but along other dimensions, *e.g.*, sample recruitment (BBJ: hospital-based, UKBB: population-based, and FinnGen: mixed), phenotyping (disease diagnosis, laboratory measurement, etc.), and environment. These other sources of heterogeneity could contribute to the differences we observe across the three biobanks. Despite these differences, we identify in this study a very substantial set of variants extremely likely to be directly causal, supported by consistency across populations, a strong enrichment of coding variants in high-PIP variants, and by the observation of 176 genes in which fine-mapping indicated multiple, independent coding variants associated with the same trait.

To our knowledge, this study provides the largest and the most comprehensive comparison of fine-mapping results from multiple large-scale biobanks of diverse ancestries. Although these data still remain limited to identify common but small-effect causal variants shared across populations, we have demonstrated that the use of diverse populations facilitates the identification of high-confidence causal variants shared across populations, population-enriched fine-mapped variants, and allelic series of high-impact variants across populations. With fast-evolving biobanks and high-throughput assays under development, our atlas of candidate causal variants provide a valuable resource for future functional characterization efforts.

### Data availability

The fine-mapping results produced by this study will be publicly available at https://www.finucanelab.org/data. The BBJ summary statistics are available at the National Bioscience Database Center (NBDC) Human Database (accession code: hum0197) and at the GWAS catalog^81^ (https://www.ebi.ac.uk/gwas/home). They are also browseable at our PheWeb^82^ website (https://pheweb.jp/). The BBJ genotype data is accessible on request at the Japanese Genotype-phenotype Archive (http://trace.ddbj.nig.ac.jp/jga/index_e.html) with accession code JGAD00000000123 and JGAS00000000114. The UKBB summary statistics will be available at the ENCODE data portal (https://www.encodeproject.org/) and at the GWAS catalog^81^ (https://www.ebi.ac.uk/gwas/home). The UKBB individual-level data is accessible on request through the UK Biobank Access Management System (https://www.ukbiobank.ac.uk/). The UKBB analysis in this study was conducted via application number 31063. The FinnGen release 6 was used in this study and is still subject to embargo according to the FinnGen consortium agreement; thus the FinnGen summary statistics are available on request (https://www.finngen.fi/en/access_results) and are being prepared for public release by Q4 2021. The GTEx v8 summary statistics is available at the GTEx Portal (https://gtexportal.org/home/datasets). The GTEx individual-level data is accessible on request through the dbGAP application (accession code: phs000424.v8.p2; https://gtexportal.org/home/protectedDataAccess). The eQTL catalogue results are available at https://www.ebi.ac.uk/eqtl/Data_access/.

### Code availability

Our fine-mapping pipeline is available at https://github.com/mkanai/finemapping-pipeline, and the code to perform all analyses and generate the figures is provided at https://github.com/mkanai/finemapping-insights. Custom fine-mapping pipelines for FinnGen is available at https://github.com/FINNGEN/finemapping-pipeline and for eQTL catalogue is available at https://github.com/eQTL-Catalogue/susie-workflow; both of which has implemented functionally-equivalent pipelines with a dataset-specific custom code.

## Data Availability

The fine-mapping results produced by this study will be publicly available at https://www.finucanelab.org/data. The BBJ summary statistics are available at the National Bioscience Database Center (NBDC) Human Database (accession code: hum0197) and at the GWAS catalog69 (https://www.ebi.ac.uk/gwas/home). They are also browsable at our PheWeb website (https://pheweb.jp/). The BBJ genotype data is accessible on request at the Japanese Genotype-phenotype Archive (http://trace.ddbj.nig.ac.jp/jga/index_e.html) with accession code JGAD00000000123 and JGAS00000000114. The UKBB summary statistics will be available at the ENCODE data portal (https://www.encodeproject.org/) and at the GWAS catalog (https://www.ebi.ac.uk/gwas/home). The UKBB individual-level data is accessible on request through the UK Biobank Access Management System (https://www.ukbiobank.ac.uk/). The UKBB analysis in this study was conducted via application number 31063. The FinnGen release 6 was used in this study and is still subject to embargo according to the FinnGen consortium agreement; thus the FinnGen summary statistics are available on request (https://www.finngen.fi/en/access_results) and are being prepared for public release by Q4 2021. The GTEx v8 summary statistics is available at the GTEx Portal (https://gtexportal.org/home/datasets). The GTEx individual-level data is accessible on request through the dbGAP application (accession code: phs000424.v8.p2; https://gtexportal.org/home/protectedDataAccess). The eQTL catalogue results are available at https://www.ebi.ac.uk/eqtl/Data_access/.

## Acknowledgements

We acknowledge all the participants of BioBank Japan, FinnGen, and UK Biobank. We thank all the members of Finucane and Daly labs for their helpful feedback. This study was supported as an ENCODE Functional Characterization Center (UM1HG009435). The BioBank Japan Project was supported by the Tailor-Made Medical Treatment program of the Ministry of Education, Culture, Sports, Science, and Technology (MEXT), the Japan Agency for Medical Research and Development (AMED). The FinnGen project is funded by two grants from Business Finland (HUS 4685/31/2016 and UH 4386/31/2016) and the following industry partners: AbbVie Inc., AstraZeneca UK Ltd, Biogen MA Inc., Celgene Corporation, Celgene International II Sàrl, Genentech Inc., Merck Sharp & Dohme Corp, Pfizer Inc., GlaxoSmithKline Intellectual Property Development Ltd., Sanofi US Services Inc., Maze Therapeutics Inc., Janssen Biotech Inc, and Novartis AG. Following biobanks are acknowledged for delivering biobank samples to FinnGen: Auria Biobank (www.auria.fi/biopankki), THL Biobank (www.thl.fi/biobank), Helsinki Biobank (www.helsinginbiopankki.fi), Biobank Borealis of Northern Finland (https://www.ppshp.fi/Tutkimus-ja-opetus/Biopankki/Pages/Biobank-Borealis-briefly-in-English.aspx), Finnish Clinical Biobank Tampere (www.tays.fi/en-US/Research_and_development/Finnish_Clinical_Biobank_Tampere), Biobank of Eastern Finland (www.ita-suomenbiopankki.fi/en), Central Finland Biobank (www.ksshp.fi/fi-FI/Potilaalle/Biopankki), Finnish Red Cross Blood Service Biobank (www.veripalvelu.fi/verenluovutus/biopankkitoiminta) and Terveystalo Biobank (www.terveystalo.com/fi/Yritystietoa/Terveystalo-Biopankki/Biopankki/). All Finnish Biobanks are members of BBMRI.fi infrastructure (www.bbmri.fi). Finnish Biobank Cooperative -FINBB (https://finbb.fi/) is the coordinator of BBMRI-ERIC operations in Finland. M.Kanai was supported by a Nakajima Foundation Fellowship and the Masason Foundation. Y.O. was supported by the Japan Society for the Promotion of Science (JSPS) KAKENHI (19H01021, 20K21834), and AMED (JP20km0405211, JP20ek0109413, JP20ek0410075, JP20gm4010006, and JP20km0405217), Takeda Science Foundation, and Bioinformatics Initiative of Osaka University Graduate School of Medicine, Osaka University. H.K.F. was funded by NIH grant DP5 OD024582 and by Eric and Wendy Schmidt.

## Methods

### Study cohorts

#### BioBank Japan (BBJ)

The BioBank Japan (BBJ) is a hospital-based cohort that collected DNA, serum, and clinical information of approximately 200,000 individuals from 66 hospitals in Japan between 2003 and 2007. All the study participants had been diagnosed with one or more of 47 target diseases by physicians at the cooperating hospitals. Written informed consent was obtained from all the participants, as approved by the ethics committees of the RIKEN Center for Integrative Medical Sciences, and the Institute of Medical Sciences, the University of Tokyo. Details of study design, sample collection, and baseline clinical information were described elsewhere^4,83^.

We genotyped samples using i) the Illumina HumanOmniExpressExome BeadChip or ii) a combination of the Illumina HumanOmniExpress and the HumanExome BeadChip. We applied standard quality-control criteria for samples and variants as described elsewhere^84^ (summarized in **Supplementary Table 1**). We analyzed 178,726 individuals of Japanese ancestry, chosen based on sample selection criteria using principal component analysis (PCA)^5^. The genotypes were prephased using Eagle^85^ and imputed using Minimac3^86^ with a reference panel that consists of the 1000 Genomes Project Phase 3 (version 5) samples (*n* = 2,504)^87^ and whole-genome sequencing (WGS) data of Japanese individuals (*n* = 1,037)^88^. We excluded variants with low imputation quality (Rsq ≤ 0.7) and used 13,531,752 variants in this study. All the variants were processed on the human genome assembly GRCh37.

We defined phenotypes based on clinical information retrieved from medical records and interviews using a standardized questionnaire. Detailed phenotype definitions are described elsewhere^5^ and summarized in **Supplementary Table 2**.

#### FinnGen

FinnGen is a public-private partnership project combining genotype data from Finnish biobanks and digital health records from Finnish health registries^6^. This study used the Data Freeze 6 which contains 271,341 individuals of Finnish ancestry. Patients and control subjects in FinnGen provided informed consent as described in **Supplementary Note**. Detailed characteristics of the cohort are described in our accompanying paper^6^.

Samples were primarily genotyped using the FinnGen ThermoFisher Axiom custom array. The samples from legacy cohorts have previously been genotyped using various generations of Illumina GWAS arrays. The genotypes were prephased using Eagle 2.3.5 and imputed using Beagle 4.1 with a reference panel of Finnish WGS data, the SISu v3 reference panel (n = 3,775). We applied post-imputation quality control as described in our accompanying paper^6^, excluding variants with INFO < 0.6 and MAF < 0.001, and used 16,311,902 variants in our study. All the variants were originally processed on the human genome assembly GRCh38, and lifted over to GRCh37 for comparison with other cohorts used in this study.

Clinical endpoints were defined based on medical records from multiple national health registries. Detailed phenotype definitions are described in our accompanying paper^6^ and summarized in **Supplementary Table 2**.

#### UK Biobank (UKBB)

The UK Biobank (UKBB) is a population-based cohort that recruited approximately 500,000 individuals in the United Kingdom between 2006 and 2010. This study analyzed a set of 366,194 unrelated “white British” individuals defined previously in the Neale Lab GWAS (https://github.com/Nealelab/UK_Biobank_GWAS). The individuals of British ancestry were determined by the PCA-based sample selection criteria (https://github.com/Nealelab/UK_Biobank_GWAS/blob/master/ukb31063_eur_selection.R), and were further filtered to self-reported “white British”, “Irish”, or “white”. The UK Biobank analysis was conducted via application number 31063. The cohort characteristics were extensively described elsewhere^7^.

Genotyping was performed using either i) the Applied Biosystems UK BiLEVE Axiom Array or ii) UKB Axiom Array. The genotypes were imputed using IMPUTE4 with a combination of reference panels: i) the Haplotype Reference Consortium and ii) UK10K and the 1000 Genomes Phase 3. We retained 13,791,467 variants with INFO > 0.8, MAF > 0.001, and Hardy-Weinberg equilibrium *P* value > 1.0 × 10^−10^, with exception for the VEP-annotated coding variants where we allowed MAF > 1.0 × 10^−6^. The detailed quality-control criteria were described in the Neale Lab GWAS (https://github.com/Nealelab/UK_Biobank_GWAS). All the variants were processed on the human genome assembly GRCh37.

We derived phenotypes based on multiple data sources available in UKBB, *e.g.*, biomarkers, body measures, and disease case-control status mapped on phecodes^89^ (https://phewascatalog.org/phecodes). Detailed phenotype definitions are described in our accompanying paper^8^ and summarized in **Supplementary Table 2**.

### Genome-wide association analysis

We performed GWAS using a generalized linear mixed model as implemented in SAIGE^36^ (for binary traits) or BOLT-LMM^37,38^ (for quantitative traits) with age, sex, top principal components, and other study-specific covariates as detailed in **Supplementary Table 1**. We excluded sex-adjusting covariates from sex-specific or stratified traits (*i.e.*, age at menarche/menopause, breast cancer, testosterone levels, and uterine fibroid; **Supplementary Table 2**). For mosaic loss of chromosome Y, we used summary statistics publicly available from BBJ^90^ and UKBB^91^.

### Statistical fine-mapping

We conducted statistical fine-mapping using FINEMAP^14,15^ and SuSiE^16^ with GWAS summary statistics and in-sample dosage LD. We defined fine-mapping regions based on a 3 Mb window around each lead variant and merged regions if they overlapped. We excluded the major histocompatibility complex (MHC) region (chr 6: 25–36 Mb) from analysis due to extensive LD structure in the region. Allowing up to 10 causal variants per region, we derived up to 10 independent 95% credible sets (CS) and posterior inclusion probabilities (PIP) of each variant using the default uniform prior probability of causality. The 95% CS reported by FINEMAP and SuSiE each have 95% posterior probability of containing a causal variant; in a locus with multiple causal variants identified, there will be multiple CS. This definition of CS differs from the definition given in Hormozdiari et al.^92^, in which each CS has 95% posterior probability of containing all causal variants in a locus. We computed in-sample dosage LD using LDstore 2 (ref. ^93^).

We combined fine-mapping results from the two methods by taking an average of PIP, excluding variants with a substantial PIP difference (> 5%) to further improve fine-mapping accuracy. We justify our approach based on functional enrichment analysis that demonstrates that the variants with inconsistent PIP across the methods show little functional enrichment (as described in our accompanying paper^8^). If either fine-mapping method failed (*e.g.*, due to conversion failure or available memory restrictions), we used successful results from the other method. If both of the methods failed, we excluded these regions from analysis.

To define independent CS merged across populations, we merged SuSiE 95% CS from each population using hierarchical clustering based on the weighted Jaccard similarity index. Briefly, we computed the PIP-weighted Jaccard similarity index between all the pairs of CS for the same trait identified from each cohort. For a pair of CS, we computed the similarity index as ∑*_i_ min*(*x_i_*, *y_i_*)/ ∑*_i_ max*(*x_i_*, *y_i_*) where *x*_i_ and *y*_i_ are PIP values (or zero if missing) in each CS for the same variant *i*. We then used 1 – the similarity index as a distance to conduct hierarchical clustering of the CS using the complete linkage method. We cut a dendrogram tree at a height of 0.9 so that any two credible sets with PIP-weighted Jaccard similarity above 0.1 are merged into a single credible set.

### Colocalization

We conducted colocalization of fine-mapped variants from complex trait and *cis*-eQTL associations. Based on fine-mapping results from complex trait and *cis*-eQTL, we computed a posterior inclusion probability of colocalization for a variant as a product of PIP for GWAS and for *cis*-eQTL (PIP_coloc_ = PIP_GWAS_ × PIP*_cis_*_-eQTL_)^92^. We assembled fine-mapping results of *cis*-eQTL associations from GTEx^47^ v8 (detailed in our accompanying paper^8^) and eQTL catalogue^48^ release 4, both of which used the same or the functionally-equivalent fine-mapping pipelines to our GWAS fine-mapping (see **Code availability**). All the variants were originally processed on the human genome assembly GRCh38 and lifted over to GRCh37 to colocalize with GWAS results in this study.

### Functional enrichment

We performed functional enrichment analysis for fine-mapped variants from each population. We first defined seven main distinct functional categories: pLoF (predicted loss-of-function), missense, synonymous, 5’ UTR, 3’ UTR, promoter, cis-regulatory element (CRE), and non-genic. We assign fine-mapped variants to these categories in the sequential order so that each category is non-overlapping from each other. Variant-based categories (pLoF, missense, synonymous, and 5’/3’ UTR variants) are defined based on the most severe consequence for a variant on a canonical transcript, predicted by the Ensembl Variant Effect Predictor (VEP)^94^ v85 (using GRCh37 and GENCODE v19). The pLoF category represents stop-gained, splice site disrupting, and frameshift variants predicted as high-confidence by LOFTEE^69^. The missense category includes missense-like variants such as low-confidence LoF. Region-based categories (promoter and CRE) are defined using region-based annotations. The promoter annotation is retrieved from the baseline annotations in Finucane & Bulik-Sullivan *et al.*^40^, originally from the UCSC Genome Browser^95^ and post-processed by Gusev *et al*^96^. The CRE annotation is defined as intersection of DNase I hypersensitive sites (DHS) and H3K27ac regions from the Roadmap Epigenomics Project^97^, ChIP-Atlas^98^, Meuleman *et al.*^99^, Domcke, *et al.*^100^, Corces *et al.*^101^, Buenrostro, *et al.*^102^, and Calderon, *et al.*^103^, reprocessed in our accompanying paper^8^. Lastly, the non-genic category represents any variants that do not belong to the other six categories. In addition, we annotated each variant using 35 binary annotations from the baselineLD v2.2 model^43^.

For each variant, we computed the maximum PIP across traits in BBJ, FinnGen, UKBB, and all cohorts combined. We estimated functional enrichment for each category as a relative risk (*i.e.*, a ratio of proportion of variants) between being in an annotation and fine-mapped (PIP ≤ 0.01 or PIP > 0.9). That is, a relative risk = (proportion of variants with PIP > 0.9 that are in the annotation) / (proportion of variants with PIP ≤ 0.01 that are in the annotation). The 95% confidence intervals are calculated using bootstrapping with 5,000 replicates.

### Fine-mapping replication analysis

To investigate fine-mapping replication, we systematically evaluated the consistency of fine-mapping results across populations for the 26 traits analyzed in all three populations (**Supplementary Table 2**), using all six pairs of discovery population and distinct secondary population. Starting from high-PIP (> 0.9) variant-trait pairs in the discovery population, we first split them by whether the association is genome-wide significant (*P* < 5.0 × 10^−8^) in the secondary population, and then categorized each pair into the following categories, based on criteria evaluated in the secondary population:

For genome-wide significant (*P* < 5.0 × 10^−8^) variant-trait pairs,

1. Pairs for which the fine-mapping result is replicated (PIP > 0.1).
2. Pairs for which the fine-mapping is not replicated (PIP ≤ 0.1) For non-genome-wide significant (*P* ≥ 5.0 × 10^−8^) variant-trait pairs,
3. Pairs for which the association is replicated (*P* < 0.01).
4. Pairs for which the association is not replicated (*P* ≥ 0.01) but the variant is included in the study and has decent statistical power (estimated power > 0.9 for achieving *P* = 0.01). We estimated statistical power via the non-centrality parameter (NCP) of the chi-square distribution^104^. We defined NCP = 2*f*(1 – *f*)*nβ*^2^ where *f* is MAF, *n* is the effective sample size, and *β* is a posterior effect size estimated by SuSiE in the discovery population. Here, we assumed the variant has the same causal effect size in a second population. For quantitative traits, effective sample size equals the number of samples. For binary traits, effective sample size is calculated via *nϕ*(1 – *ϕ*) where *ϕ* is the number of cases divided by the number of total samples. We note that this power estimation does not account for linear mixed models adopted by BOLT-LMM or SAIGE.
5. Whether the variant is analyzed in the study (*i.e.*, exists in summary statistics). The missingness is mainly due to low frequency or monomorphism (non-existence) in the secondary population, which is described in the **Supplementary Note**.

The schematic flowchart of this process is illustrated in **Fig. 2a**. We note that there could be a case where non-genome-wide significant variant-trait pairs (*P* ≥ 5.0 × 10^−8^) in a secondary population still had fine-mapping replication (PIP > 0.1).

### High-confidence and low-confidence fine-mapping results

We annotated high-confidence and low-confidence high-PIP (> 0.9) variant-trait pairs for the 91 traits analyzed in two or more populations (**Supplementary Table 3**). High-confidence pairs are defined as having PIP > 0.9 in at least one population and PIP > 0.1 in all the other populations analyzed in this study. Low confidence pairs are defined as having PIP > 0.9 in one population and *P* < 5.0 × 10^−8^ but PIP ≤ 0.1 and not in 95% CS in one of the other populations. Those categorized otherwise (*e.g.*, population-specific variants) were not assigned either annotation.

### Allele frequency enrichment

To identify population-enriched variants, we defined allele frequency (AF) enrichment metrics as a ratio of pseudo AF between ancestral and founder populations. To do this, we retrieved allele counts from gnomAD^69^ v2 and GEM-J WGS^70^. To account for finite sample sizes, we computed pseudo AF by constantly adding one to allele count (AC), *i.e.*, pseudo AF = (AC + 1) / allele number. Due to the disparity in available sample sizes between gnomAD v2 exomes and genomes, we computed enrichment metrics separately for coding and non-coding variants using exomes and genomes, respectively. Coding and non-coding variants are defined as having VEP-predicted coding consequences or not (see the previous section).

For coding variants, we used gnomAD v2 exomes for the Finnish (*n* = 10,824), non-Finnish-Swedish-Estonian Europeans (NFSEE; *n* = 43,697), and non-Japanese-Korean East Asians (NJKEA; *n* = 7,212). For non-coding variants, we used gnomAD v2 genomes for the Finnish (*n* = 1,738), NFSEE (*n* = 5,421), and NJKEA (*n* = 780). We used the GEM-J WGS for both coding and non-coding variants, which contains WGS data from the Japanese population (*n* = 7,609). To account for coverage differences across data sources, we excluded regions from GEM-J WGS with a median coverage < 10 in gnomAD exomes or genomes. To eliminate non-coding enrichment due to tagging coding variants, we excluded non-coding variants in LD (*r*^2^ > 0.1) with coding variants using gnomAD v2 LD matrices for the Finnish and East Asian populations. We restricted our analysis to 140,416 and 91,564 coding variants and 11,732,074 and 9,539,454 non-coding variants tested in FinnGen and BBJ GWAS, respectively. To annotate estimated allele age, we retrieved point estimates of allele age (mode of the composite posterior distribution) from the Genealogical Estimation of Variant Age (GEVA)^74^.

### Allelic series analysis

We investigated an allelic series of fine-mapped variants within and across populations. We first took nonsynonymous coding variants (pLoF and missense predicted by VEP as described in the previous section) that had PIP > 0.1 for at least one of the studied traits. We then counted the number of these variants falling in each gene, identified allelic series of two or more such variants in a single gene for the same trait, and categorized allelic series according to whether they were discoverable in a single population or only by combining data across populations. Furthermore, we investigated non-coding variants that are proximal to these fine-mapped nonsynonymous coding variants (< 100 kb), assuming they might act through the same gene.

## Extended Data Figures

**Extended Data Fig. 1.**
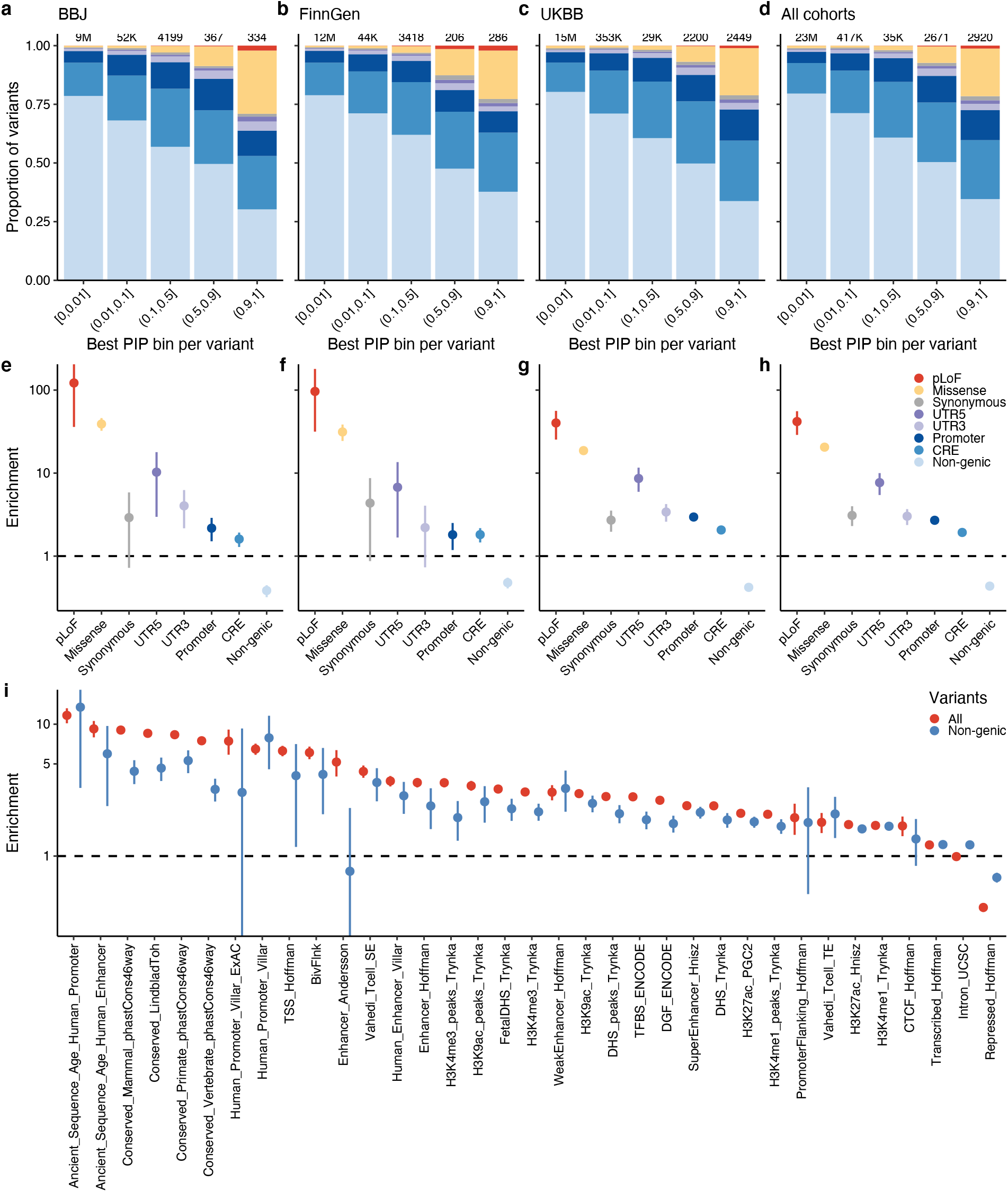
Functional enrichments of fine-mapped variants. **a–d.** Proportion of variants for the seven main functional categories (**Methods**), stratified by the best PIP bin for a variant in BBJ, FinnGen, UKBB, and all cohorts combined. Labels above each bar represent the number of variants in each bin. **e-h**. Enrichments of fine-mapped variants (PIP > 0.9) in each functional category compared to non-fine-mapped variants (PIP ≤ 0.01). **i**. Enrichments in 35 binary annotations from the baselineLD v2.2 model^43^. Enrichment was calculated as a relative risk (*i.e.*, a ratio of proportion of variants) between being in an annotation and fine-mapped (PIP ≤ 0.01 or PIP > 0.9; **Methods**). Error bars correspond to 95% confidence intervals using bootstrapping. Numerical results are available in **Supplementary Table 5,6**.

**Extended Data Fig. 2.**
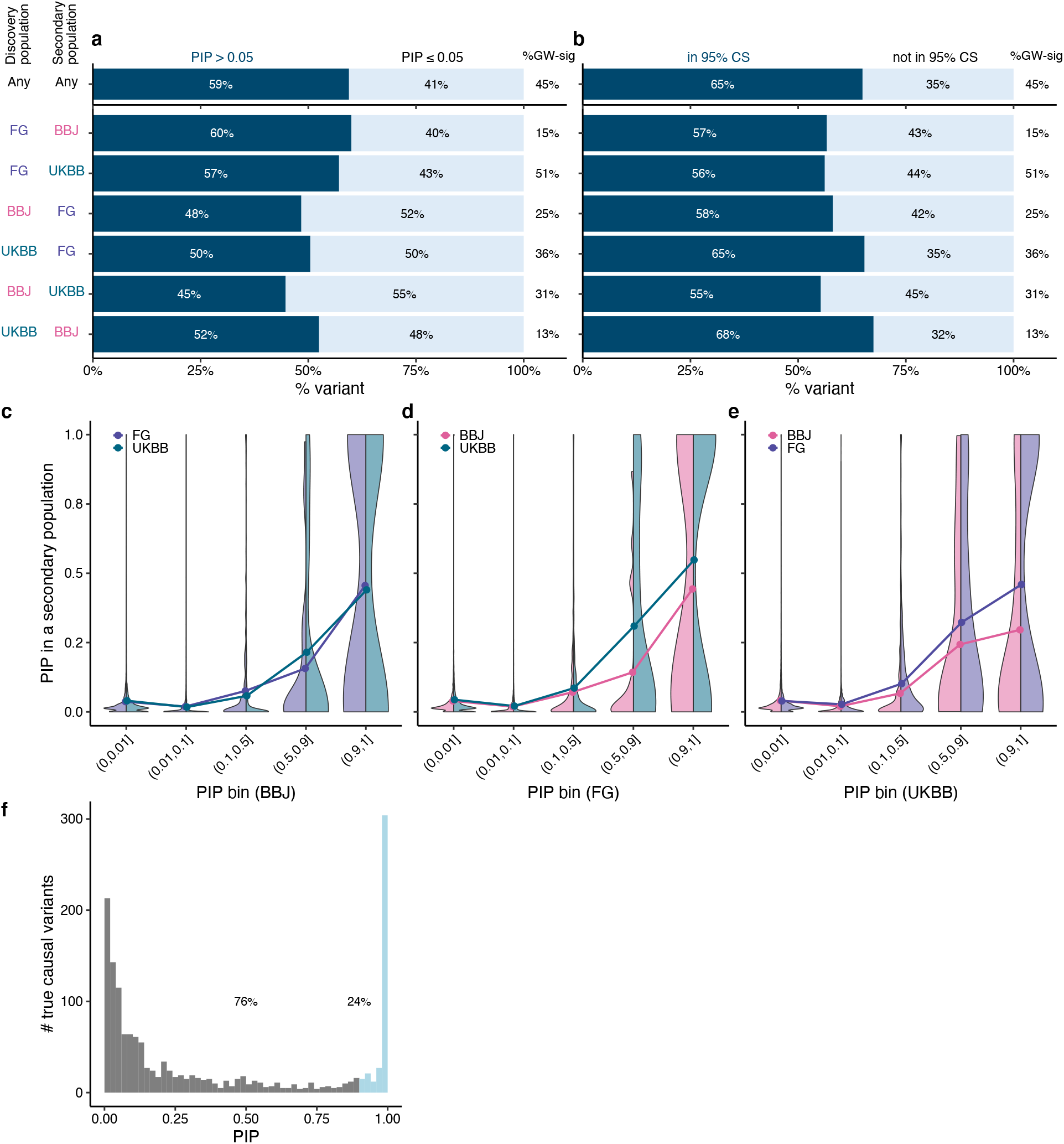
Additional details of fine-mapping replication status across populations. **a,b**. Breakdowns for the genome-wide significant variant-trait pairs (*P*GWAS < 5.0 × 10^−8^) in a secondary population, using distinct fine-mapping replication criteria (**a**. PIP > 0.05 and **b**. in 95% CS) different from Fig. 2 (PIP > 0.1). **c–e**. PIP distributions in a secondary population, stratified by PIP bins in a discovery population. Half-sided violin plots represent PIP distributions for each secondary population. Points represent mean PIP in the secondary population for each PIP bin in a discovery population. **f**. PIP distribution of true causal variants with *P*GWAS < 5.0 × 10^−8^ in simulated GWAS data from our companion paper^8^.

**Extended Data Fig. 3.**
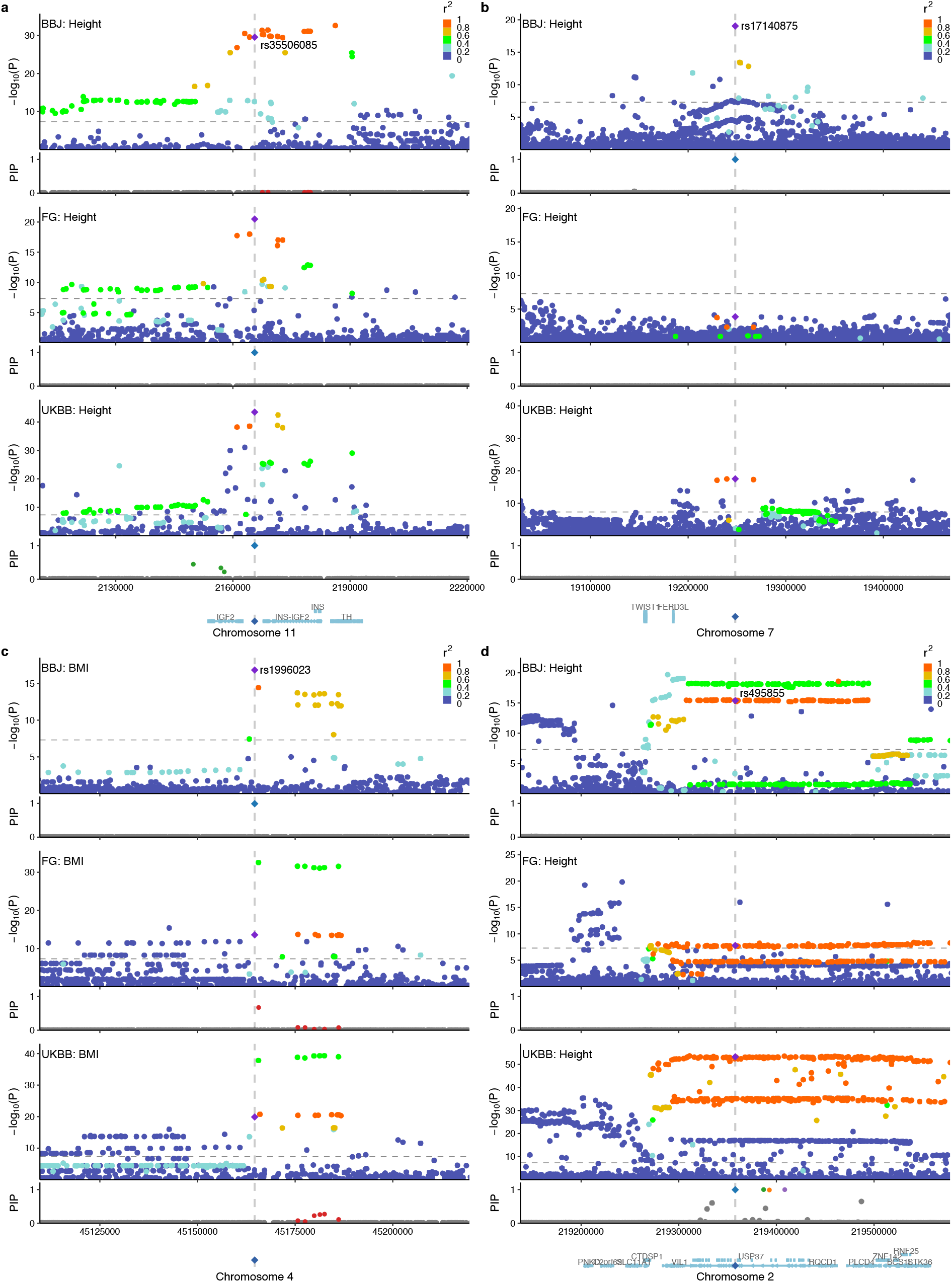
Illustrative examples of fine-mapping non-replication across populations. Locuszoom plots for the same locus across populations. Colors in the locuszoom panels represent *r*^2^ values to the lead variant. In the PIP panels, only fine-mapped variants in SuSiE 95% CS are colored, where the same colors are applied across populations based on the merged CS (**Methods**). **a**. rs35506085 for height that was fine-mapped in FinnGen and UKBB (PIP = 1.0), but not in BBJ (PIP ∼ 0) likely due to extensive LD. **b**. rs17140875 for height that was fine-mapped in BBJ (PIP = 1.0) but not in FinnGen or UKBB (PIP ∼ 0). The variant is more common in BBJ (MAF = 0.08) than in FinnGen or UKBB (MAF = 0.04 and 0.05, respectively) and has more LD neighbors in Europeans. **c**. rs1996023 for BMI that was fine-mapped in BBJ (PIP = 0.99), but not in FinnGen or UKBB (PIP ∼ 0). Instead, we found other CS in FinnGen and UKBB that showed modest LD with rs1996023 in Europeans (*r*^2^ ∼ 0.5) but high LD in BBJ (*r*^2^ ∼ 0.8). **d**. rs495855 for height that was fine-mapped only in UKBB (PIP = 1.0). This seems very likely a false positive given extensive LD in every population.

**Extended Data Fig. 4.**
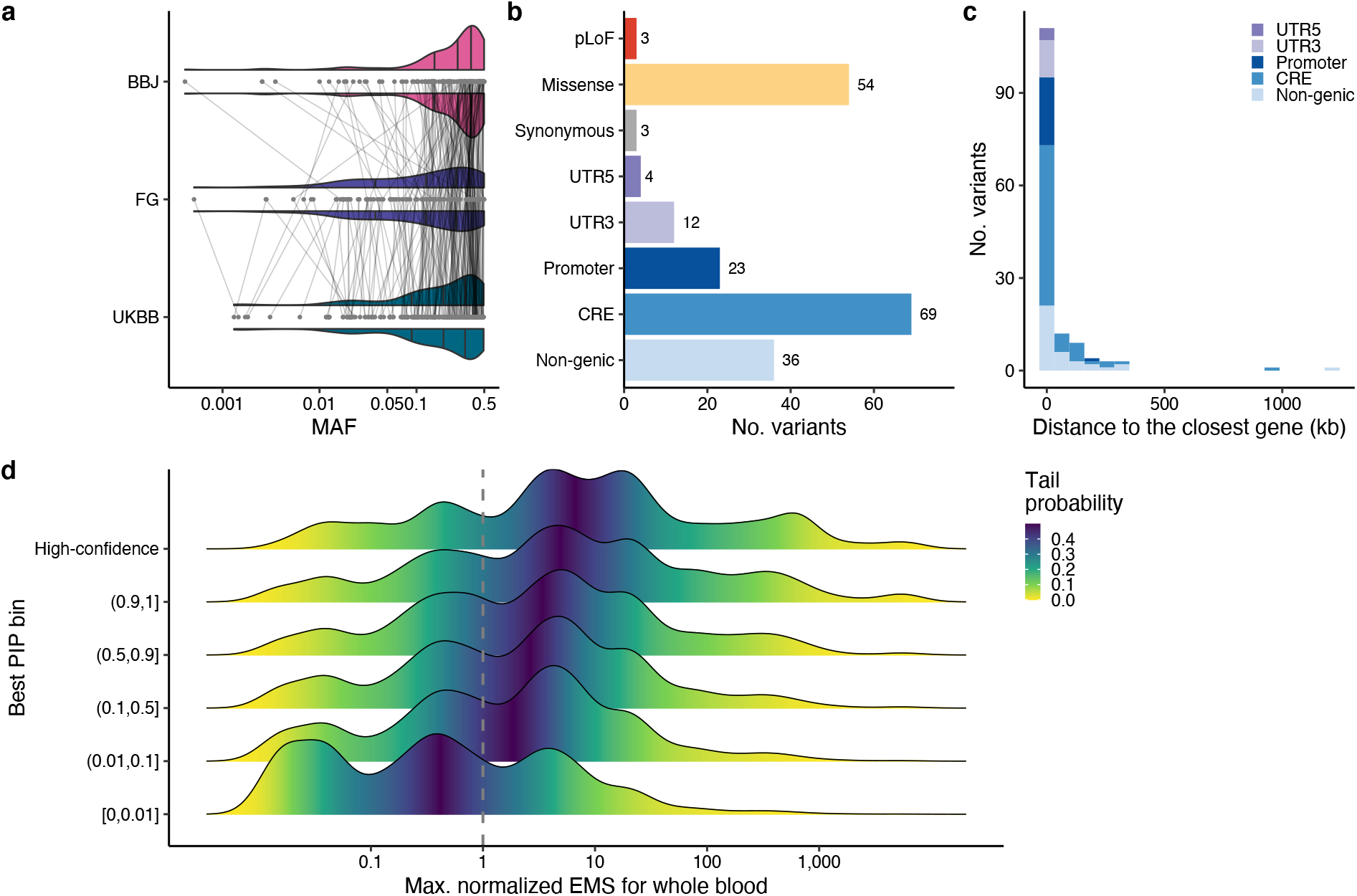
Overview of high-confidence fine-mapped variants. **a.** Distribution of minor allele frequencies (MAF) in each cohort. Violin plots represent the distribution. Each point represents a high-confidence fine-mapped variant and each line connects the same variant across cohorts. **b**. Consequences annotated by VEP (see **Methods**). **c**. Histogram of distance to the closest gene for high-confidence fine-mapped non-coding variants. Color represents non-coding consequences same as **b**. **d**. Distribution of predicted expression modifier score (EMS)^57^ for fine-mapped non-coding variants, stratified by the best PIP bins. The highest bin (0.9 < PIP ≤ 1) was further stratified into the high-confidence variants or not based on replication across populations (see **Methods**). Maximum normalized EMS score over genes was calculated for each fine-mapped variant using the whole blood tissue. Details of the high-confidence fine-mapped variants are summarized in **Supplementary Table 8,9**.

**Extended Data Fig. 5.**
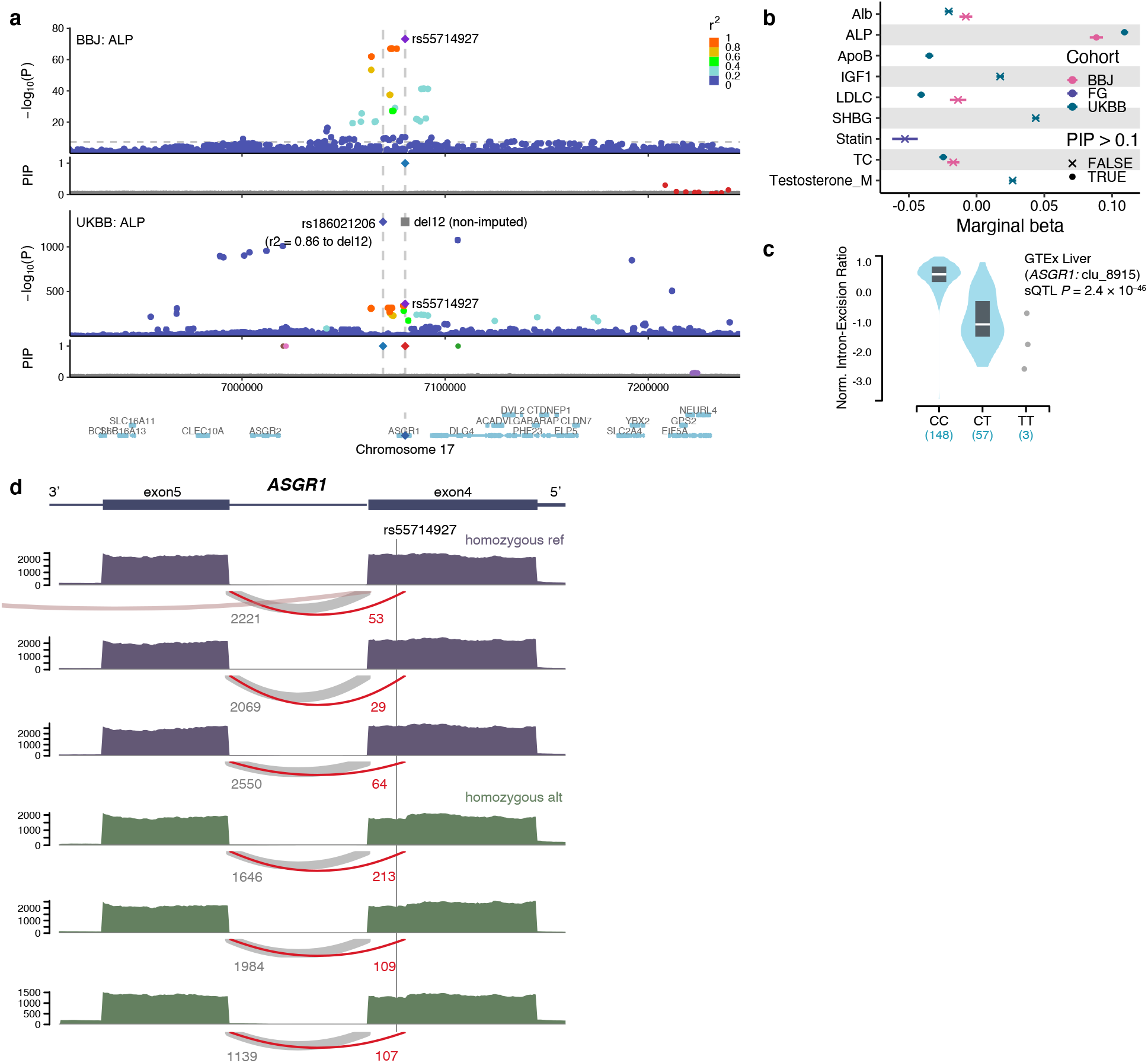
Synonymous variant rs55714927 shows splicing effect in *ASGR1*. **a.** Locuszoom plots for alkaline phosphatase (ALP) in BBJ and UKBB. **b**. Phenome-wide association study (PheWAS) of rs55714927 across all the traits analyzed in this study. Only phenotypes that showed *P* < 5.0 × 10^−8^ in any cohort are displayed. Each point represents a marginal beta for a given trait in a cohort, with an error bar representing the standard error. Shape of each point represents whether each variant showed PIP > 0.1. **c**. sQTL effect of rs55714927 in GTEx liver. **d**. Sashimi plot showing splicing effects of rs55714927 in three homozygous reference allele carriers vs. three homozygous alternative allele carriers that were randomly chosen.

**Extended Data Fig. 6.**
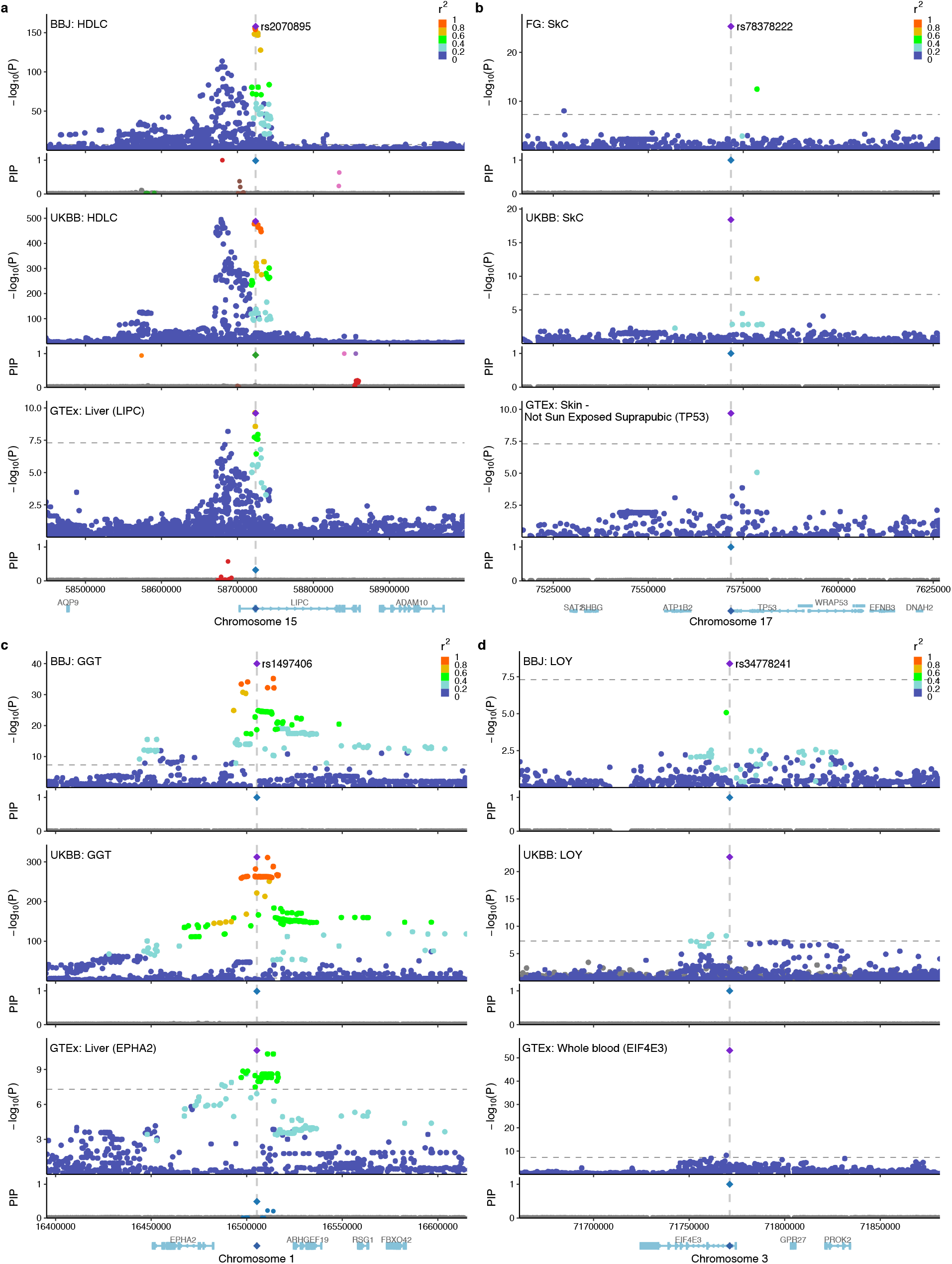
Colocalization between high-confidence fine-mapped non-coding variants for complex traits and *cis*-eQTL associations in trait-relevant tissues. Locuszoom plots for the same locus of complex traits across populations and of *cis*-eQTL associations in trait-relevant tissues. Colors in the locuszoom panels represent *r*^2^ values to the lead variant. In the PIP panels, only fine-mapped variants in SuSiE 95% CS are colored, where the same colors are applied across populations based on the merged CS (**Methods**). **a**. rs2070895 for HDL cholesterol in BBJ and UKBB and for *LIPC* expression in GTEx liver. **b**. rs78378222 for skin cancer in FinnGen and UKBB and for *TP53* expression in GTEx skin. **c**. rs1497406 for γ-glutamyl transferase (GGT) in BBJ and UKBB and for *EPHA2* expression in GTEx liver. d. rs34778241 for loss of chromosome Y (LOY) in BBJ and UKBB and for *EIF4E3* expression in GTEx whole blood.

**Extended Data Fig. 7.**
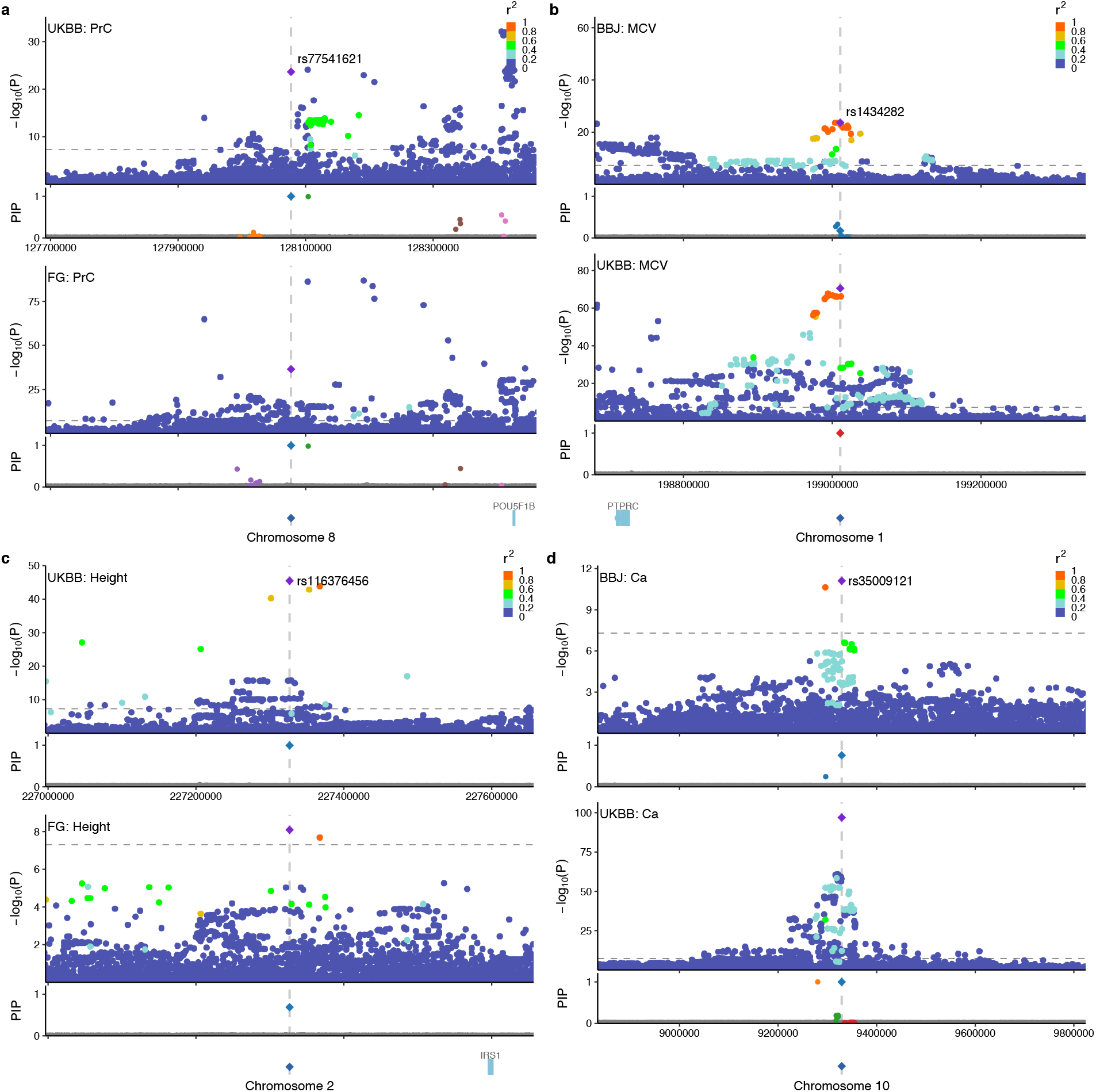
High-confidence fine-mapped intergeneric variants in a gene desert. Locuszoom plots for the same locus across populations. Colors in the locuszoom panels represent *r*^2^ values to the lead variant. In the PIP panels, only fine-mapped variants in SuSiE 95% CS are colored, where the same colors are applied across populations based on the merged CS (**Methods**). **a**. rs77541621 in the 8q24 locus for prostate cancer in UKBB and FinnGen. **b**. rs1434282 in the 1q32 locus for mean corpuscular volume (MCV) in BBJ and UKBB. **c**. rs116376456 in the 2q36 locus for height in UKBB and FinnGen. **d**. rs35009121 in the 10p14 locus for calcium levels in BBJ and UKBB.

**Extended Data Fig. 8.**
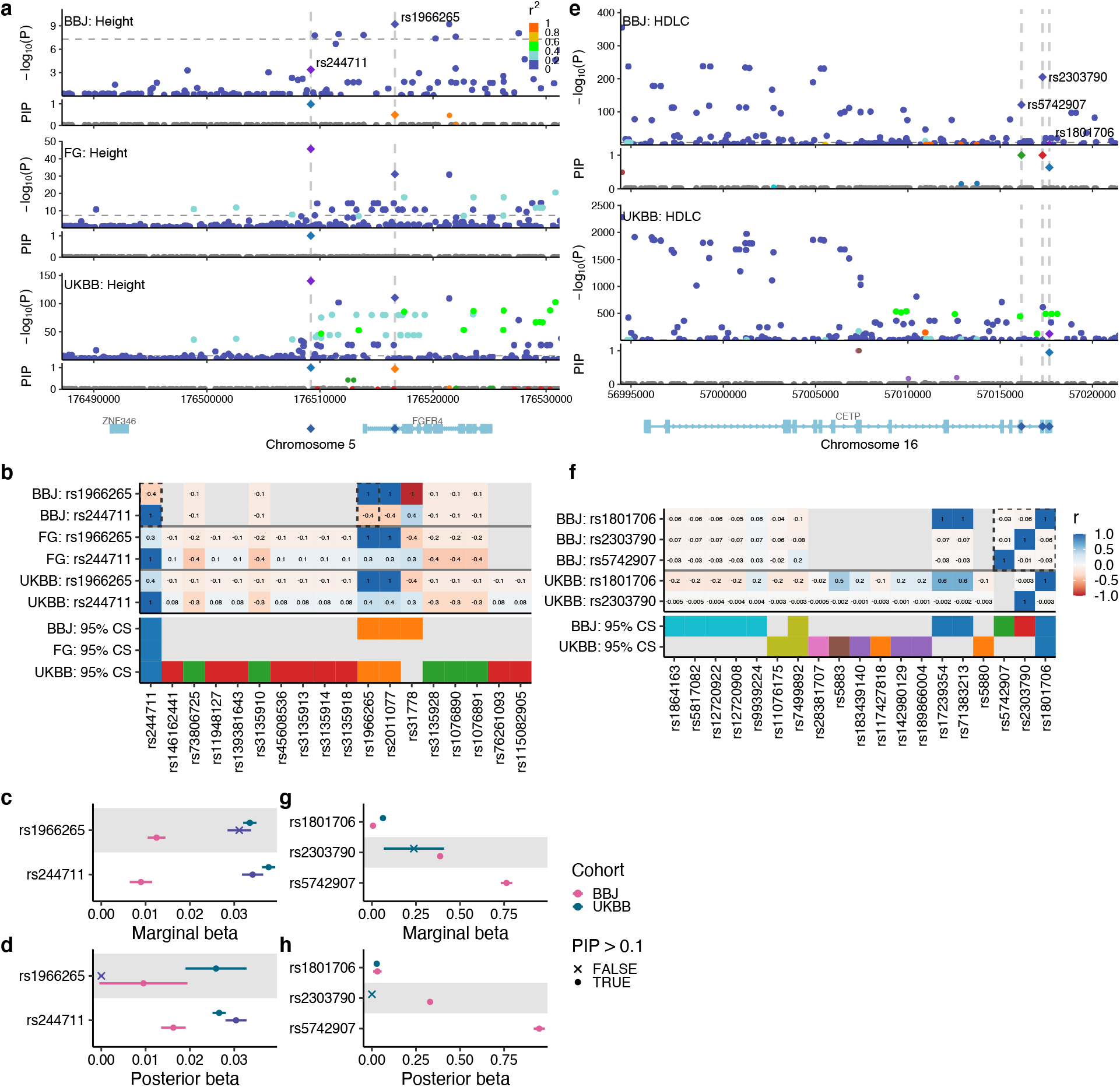
Putative causal variants are negatively correlated in a locus. **a–d.** rs244711 and rs1966265 for height in BBJ, FinnGen, and UKBB. **e–h**. rs1801706, rs5742907 and rs2303790 for HDL cholesterol in BBJ and UKBB. **a, e.** Locuszoom plots for the same locus across populations. Colors in the manhattan panels represent *r*^2^ values to the lead variant. In the PIP panels, only fine-mapped variants in SuSiE 95% CS are colored, where the same colors are applied across populations based on the merged CS (**Methods**). **b, f.** Heatmaps showing *r* values between the highlighted variants and the other variants in 95% CS in each population. In a CS panel, variants are colored by the same colors in the locuszoom plots (**a, e)**. **c, d, g, h**. Forest plots showing marginal and posterior betas of fine-mapped variants. Point color represents each cohort and shape represents whether the variant showed PIP > 0.1 in each cohort.

**Extended Data Fig. 9.**
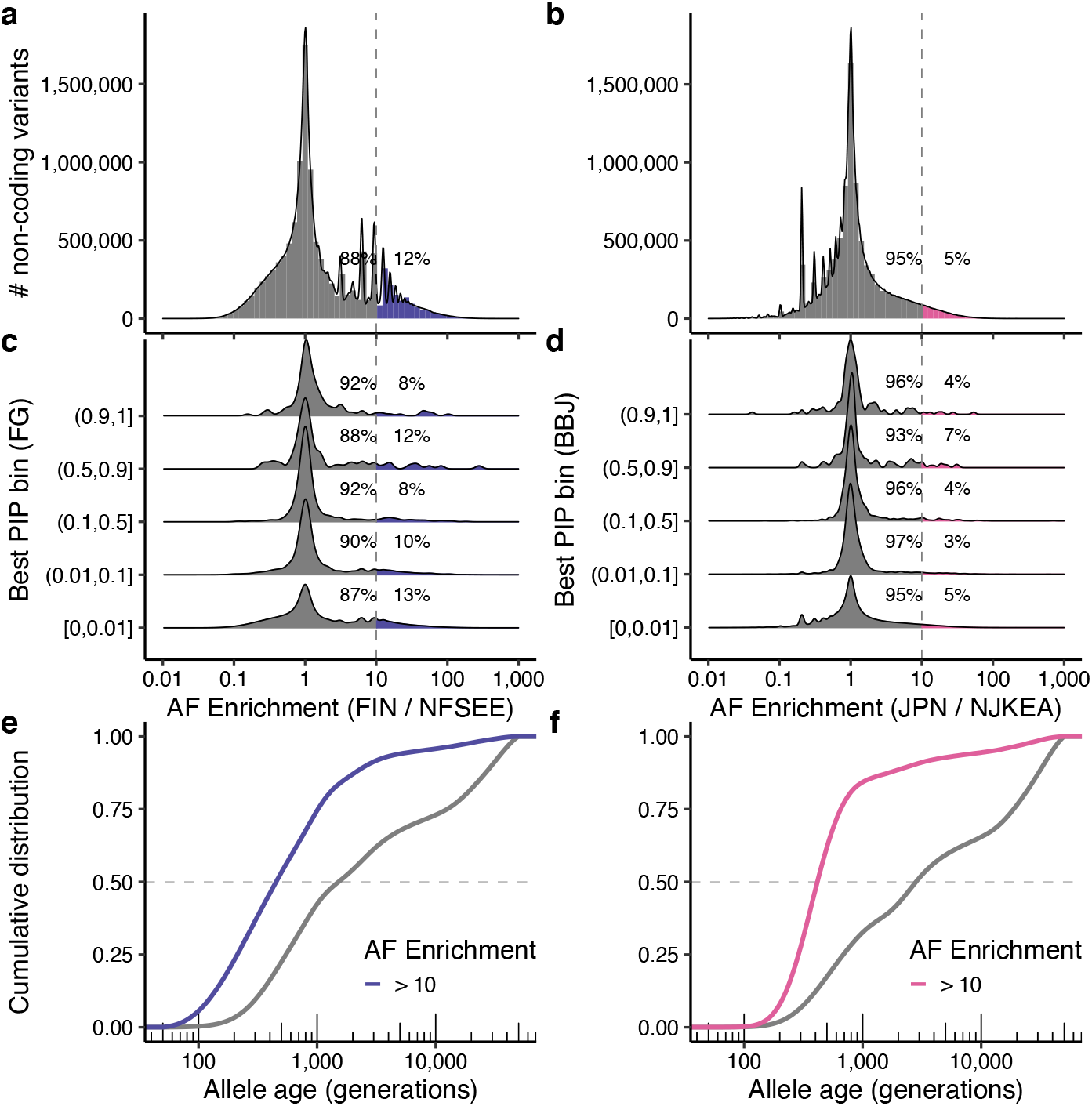
Population-enriched non-coding variants. **a–d.** Histograms showing a distribution of allele frequency (AF) enrichment metric in (**a**) Finnish (*n* = 1,738) and (**b**) Japanese (*n* = 7,609) populations. A ratio of AF was computed against NFSEE (*n* = 5,421) and NJKEA (*n* = 780) for non-coding variants analyzed in FinnGen or BBJ GWAS that exist in gnomAD WGS or GEM-J WGS, respectively. For a subset of variants that are fine-mapped in our analysis (see **Methods**), we show AF enrichment distribution across maximum PIP bins computed in (**c**) FinnGen or (**d**) BBJ. **e–f**. Cumulative distribution of estimated allele age for non-coding variants, stratified by AF enrichment in (**e**) Finnish or (**f**) Japanese. FIN: Finnish, JPN: Japanese, NFSEE: Non-Finnish-Swedish-Estonian European, NJKEA: Non-Japanese-Korean East Asian.

**Extended Data Fig. 10.**
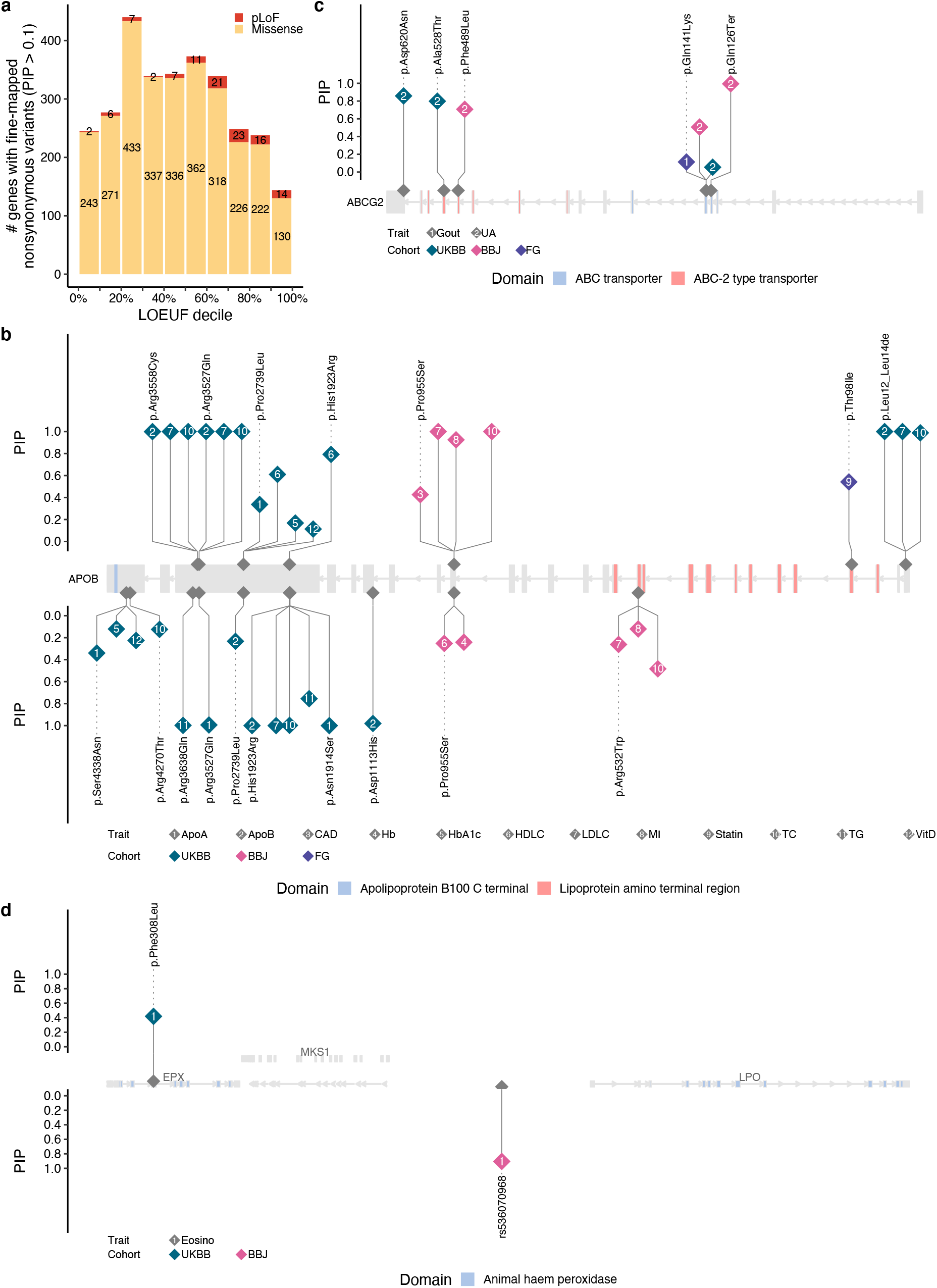
Allelic series of putative causal variants across populations. **a.** Number of genes with fine-mapped nonsynonymous variants (pLoF and missense) with best PIP > 0.1 for each LOEUF decile^69^. Genes without fine-mapped nonsynonymous variants are not plotted. Colors represent the consequence of each variant. When multiple nonsynonymous variants are found, the most deleterious consequence is colored. **b–d**. Lollipop plots of allelic series for (**b**) *APOB*, (**c**) *ABCG2*, and (**d**) *EPX*. Each point represents a fine-mapped variant from a single trait and cohort. Point color represents discovery cohort and number label represents a fine-mapped trait. Points above the gene body correspond to those with positive effect sizes, whereas points below the gene body correspond to those with negative effect sizes. Coding variants are labeled with the HGVS protein nomenclature and non-coding variants (in **d**) are labeled with rsids. Protein domains are annotated based on the Pfam database

