## Supplementary Information for "Insights from complex trait fine-mapping across diverse populations"

Kanai, M. *et al*.

### Supplementary Box

#### Novel genes implicated by population-enriched variants

Fine-mapping of complex traits in FinnGen led to the identification of Finnish-enriched missense variants in genes novel for several traits. Many of these genes possess compelling biological rationales linking them to these traits.

*THBS3*

On chromosome 1, rs199935580 corresponds to an Arginine to Tryptophan substitution (p.Arg520Trp) in the gene *THBS3*. This rare missense variant (MAF = $1.0 \times 10^{-3}$ in gnomAD Finnish; $1.2 \times 10^{-5}$ in gnomAD Non-Finnish-Swedish-Estonian Europeans [NFSEE]) is fine-mapped for increased risk of carpal tunnel syndrome ($P = 7.4 \times 10^{-10}$; $\beta = 1.44$; PIP = 1.0). In carpal tunnel syndrome (CTS), the median nerve is pinched as it traverses through the wrist. A previous GWAS on CTS revealed an important role for the extracellular matrix in its etiology[1]. *THBS3* encodes a member of the thrombospondin family, a group of proteins known for binding to various extracellular matrix proteins[2]. No specific role for *THBS3* in CTS has been noted previously. However, its closest homolog, *COMP*, is causal for familial carpal tunnel syndrome 2 (ref. [3]).

*LUM*

On chromosome 12, the rare variant rs191692991 corresponds to an Arginine to Cysteine substitution (p.Arg310Cys) in the gene *LUM*. This missense variant (MAF = $5.3 \times 10^{-3}$ in gnomAD Finnish; $1.2 \times 10^{-5}$ in gnomAD NFSEE) is strongly associated with "fibroblast disorders" such as Dupuytren's contracture, a condition in which the skin under the palm of the hand becomes thick and fibrous ($P = 6.9 \times 10^{-9}$; $\beta = 1.02$; PIP = 1.0). *LUM* encodes lumican, a leucine-rich repeat glycoprotein important for regulation of collagen fibril formation[4]. One hypothesis is that this mutation reduces stability or activity of lumican, leading to misregulation of collagen and accumulation of fibrils in the hand and elsewhere.

*POF1B*

On the X chromosome, the rare variant rs200939713 corresponds to an Arginine to Tryptophan substitution (p.Arg339Trp) in *POF1B* (MAF = $1.6 \times 10^{-3}$ in gnomAD Finnish; monomorphic in gnomAD NFSEE). This missense variant is associated with risk of varicose veins ($P = 3.4 \times 10^{-11}$; $\beta = 0.84$; PIP = 0.99), a condition in which veins just below the skin become enlarged and prominent. *POF1B* encodes a protein important for epithelial structural integrity through desmosomes[5]. It's possible this mutation reduces *POF1B* stability, reducing epidermal integrity increasing occurrence or appearance of varicose veins.

### Supplementary Note

#### "Missing" variants from summary statistics in other populations

After restricting to the 26 traits available in every population (BBJ, FinnGen, and UKBB), we found 301 high-PIP variants (PIP > 0.9) fine-mapped in a discovery population that are missing from summary statistics in other populations (**Fig. 2a–c**). We characterized reasons for the missingness using the following criteria:

1. Variants do not exist in imputation reference panels used in each cohort, *i.e.*, BBJ[6]: the 1000 Genomes Phase 3 ($n$ = 2,504) + Japanese WGS ($n$ = 1,037); FinnGen: Finnish WGS ($n$ = 3,775); and UKBB[7]: the Haplotype Reference Consortium ($n$ = 64,976) + the 1000 Genomes Phase 3 + UK10K ($n$ = 3,781).
2. Low MAF (MAF < 0.005) in a population based on the GEM-J WGS[8] for BBJ and the gnomAD[9] v2 for FinnGen and UKBB.
3. Low imputation INFO score (INFO < 0.7 for BBJ and INFO < 0.8 for FinnGen/UKBB)
4. Hardy-Weinberg equilibrium (HWE) outlier (HWE test $P$-value < $1 \times 10^{-10}$) only in UKBB.

We confirmed that the missingness are primarily due to low frequency in other populations (**Supplementary Fig. 1**). Note that since BBJ and UKBB included the 1000 Genomes Project in their reference panels, there are variants that exist in the reference but showed very low MAF in the Japanese or White British populations; This is in contrast to the FinnGen which only used a Finnish-specific reference panel.

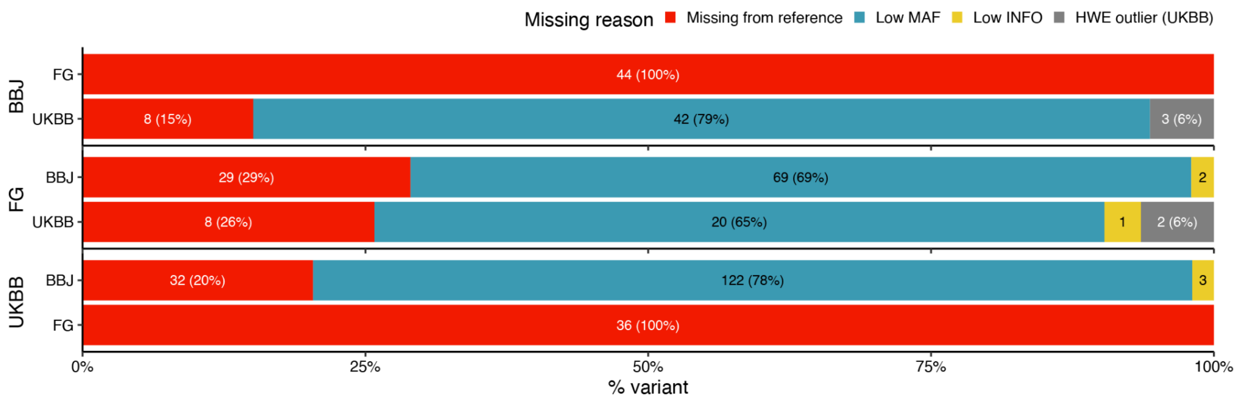


**Supplementary Fig. 1 | Overview of "missing" variants from summary statistics.** We characterized the reasons that high-PIP variants (PIP > 0.9) in a single population are missing from summary statistics in other populations. The following criteria represent imputation and quality control procedures adopted in each cohort (**Methods**). Missing from reference: Variants do not exist in imputation reference panels used in each cohort, *i.e.*, BBJ: the 1000 Genomes Phase 3 ($n$ = 2,504) + Japanese WGS ($n$ = 1,037); FinnGen: Finnish WGS ($n$ = 3,775); and UKBB: the Haplotype Reference Consortium ($n$ = 64,976) + the 1000 Genomes Phase 3 + UK10K ($n$ = 3,781). Low MAF: MAF < 0.005 in a population based on the GEM-J WGS for BBJ and the gnomAD v2 for FinnGen and UKBB. Low INFO: INFO < 0.7 for BBJ and INFO < 0.8 in FinnGen/UKBB. HWE outlier (UKBB): HWE P-value < 1e-10 (only in UKBB).

##### *Low INFO variants*

We found six high-PIP variant-trait pairs are missing from summary statistics in other populations due to low INFO score despite having a high MAF in a population:

- rs138381300 (frameshift, *FLG*:p.Ser761CysfsTer36) fine-mapped for atopic dermatitis (PIP = 1.0) in FinnGen, but missing from UKBB (INFO = 0.60 in UKBB; MAF = 0.02 in non-Finnish Europeans). This variant is monomorphic in Japanese.
- rs6874142 (intron variant of *STC2*) fine-mapped for height (PIP = 1.0) in UKBB, but missing from BBJ (INFO = 0.64 in BBJ; MAF = 0.03 in Japanese). This variant is significantly associated in FinnGen ($P = 1.6 \times 10^{-10}$) but not fine-mapped (PIP = 0.008).
- rs117137535 (intron variant of *ARRDC1*) fine-mapped for atopic dermatitis (PIP = 1.0) in FinnGen, but missing from BBJ (INFO = 0.55 in BBJ; MAF = 0.10 in Japanese). This variant is not associated in UKBB ($P = 0.75$).
- rs9893867 (intron variant of *SLC43A2*) fine-mapped for type 2 diabetes (PIP = 0.97) in FinnGen, but missing from BBJ (INFO = 0.62 in BBJ; MAF = 0.08 in Japanese). This variant is not associated in UKBB ($P = 0.15$).
- rs11653578 (intron variant of *ANKFN1*) fine-mapped for type 2 diabetes (PIP = 0.98) in UKBB, but missing from BBJ (INFO = 0.61 in BBJ; MAF = 0.23 in Japanese). This variant is not associated in FinnGen ($P = 0.20$).
- rs3810291 (3' UTR variant of *ZC3H4*) fine-mapped for body mass index (PIP = 0.96) in UKBB, but missing from BBJ (INFO = 0.61 in BBJ; MAF = 0.23 in Japanese). This variant is significantly associated in FinnGen ($P = 1.4 \times 10^{-10}$) but not fine-mapped (PIP = 0.009).

There are a few potential reasons for relatively low INFO scores of these variants. First, rs138381300 is a frameshift variant of *FLG* which is known to have a highly repetitive coding sequence. This makes short-read next-generation sequencing (NGS) extremely challenging; and indeed, the region is registered as NCBI GeT-RM NGS Dead Zone[10]. Second, we found three variants are located near the telomere regions (rs6874142: 5q35.1, rs117137535: 9q34.3, and rs9893867: 17p13.3) which are difficult to impute. Lastly, although we did not find any simple reason for rs11653578 and rs3810291, we speculate that the low INFO scores of these variants are due to a combination of several factors including reference panel and genotyping array quality, given that their INFO scores are just borderline below the threshold (INFO = 0.61 < 0.7).

Of the six pairs, we are confident that rs138381300 is a putative causal variant for atopic dermatitis (PIP = 1.0 in FinnGen) since it is a frameshift variant for the known pathogenic gene *FLG*. The rest of the variant-trait pairs show varying evidence of association, emphasizing the critical needs for replication in fine-mapping studies.

*HWE outlier variants in UKBB*

In addition, we observed that five high-PIP variant-trait pairs (three unique variants) are missing from UKBB summary statistics due to HWE outlier, namely:

- rs2237897 (intron variant of *KCNQ1*) fine-mapped for body mass index, body weight, and type 2 diabetes (PIP =1.0) in BBJ (MAF = 0.04, HWE *P*-value = $6.3 \times 10^{-134}$ in UKBB).
- rs4765138 (intergenic variant) fine-mapped for height (PIP = 0.99) in FinnGen (MAF = 0.31, HWE *P*-value = $5.7 \times 10^{-32}$ in UKBB).
- rs117952254 (intergenic variant) fine-mapped for myocardial infarction (PIP = 0.98) in FinnGen (MAF = 0.027, HWE *P*-value = $2.9 \times 10^{-44}$ in UKBB).

We previously reported that UKBB imputed data contain genotyped SNPs failing the HWE test (http://www.nealelab.is/blog/2019/9/17/genotyped-snps-in-uk-biobank-failing-hardy-weinberg-equilibrium-test). Briefly, we observed 15,069 genotyped variants that are retained in the imputed bgen files with INFO = 1 and HWE p-value < $1 \times 10^{-12}$, due to UKBB's QC criteria relying on a per-batch HWE test[7] (**Supplementary Fig. 2**). This observation was particularly concerning given that we found that 3,987 of these variants have no homozygous alternative genotypes despite having a MAF > 1%. To mitigate this issue, we applied an additional post-hoc filtering that excludes any imputed variants with HWE test *P*-value < $1 \times 10^{-10}$; however, this might exclude a potential causal variant too.

Having said that, we are confident rs2237897 is a putative causal variant (PIP = 1.0 and 0.31 in BBJ and FinnGen, respectively) that confers a risk for type 2 diabetes as previously reported[11]. However, rs4765138 and rs117952254 are not well-characterized in the current literature, with lack of fine-mapping replication in BBJ (rs4765138: *P* = $4.1 \times 10^{-19}$ and PIP = $1.1 \times 10^{-5}$ for height; rs117952254: missing in BBJ), suggesting that further replication effort should be warranted.

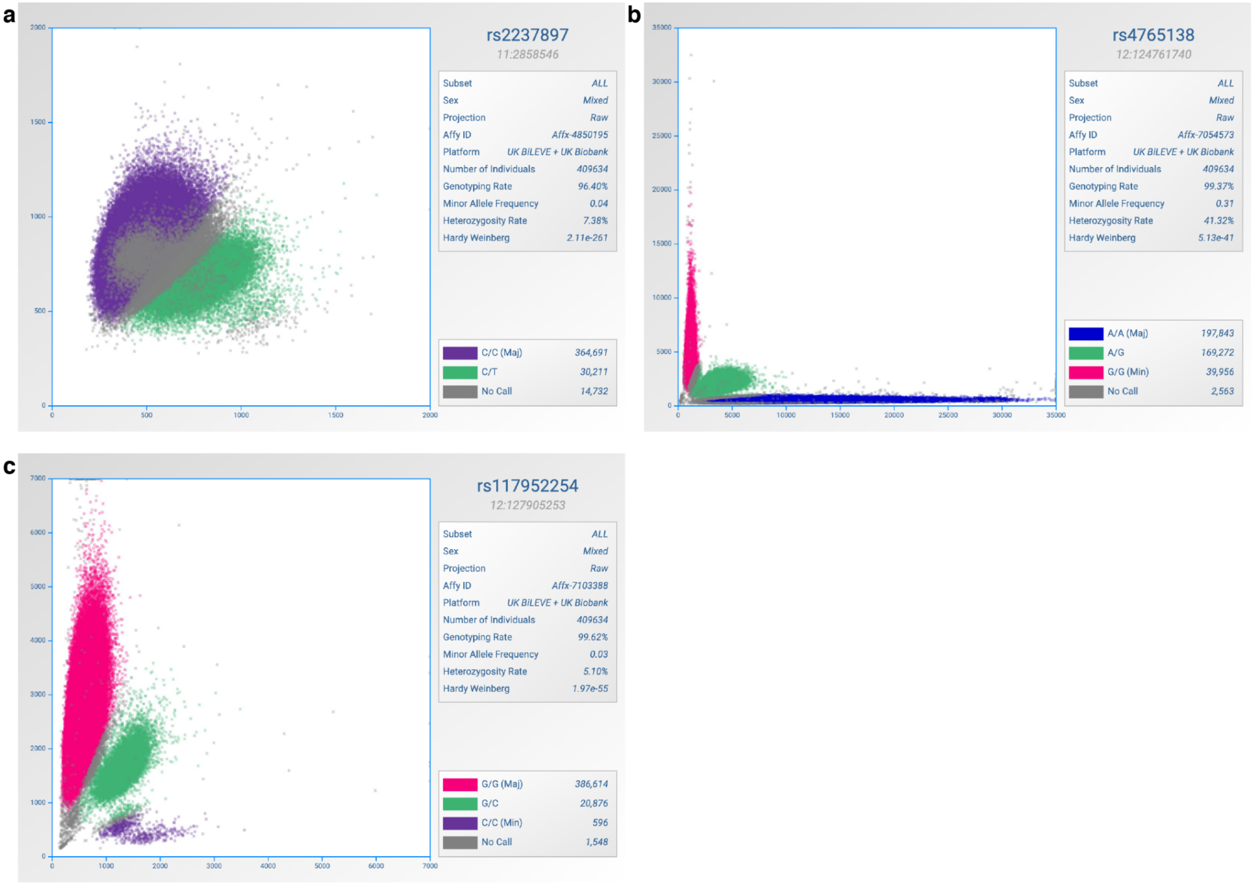


**Supplementary Fig. 2 | Genotype cluster plots in UKBB White British individuals.** Cluster plots of genotyped SNP intensity are shown for three SNPs: **a.** rs2237897, **b**. rs4765138 and **c**. rs117952254. Colors correspond to called genotypes. All the variants passed a per-batch QC but showed HWE test *P*-value < $1 \times 10^{-10}$ in aggregate. The cluster plots were generated by ScatterShot (http://mccarthy.well.ox.ac.uk/static/software/scattershot/).

### FinnGen ethics statement

Patients and control subjects in FinnGen provided informed consent for biobank research, based on the Finnish Biobank Act. Alternatively, separate research cohorts, collected prior the Finnish Biobank Act came into effect (in September 2013) and start of FinnGen (August 2017), were collected based on study-specific consents and later transferred to the Finnish biobanks after approval by Fimea, the National Supervisory Authority for Welfare and Health. Recruitment protocols followed the biobank protocols approved by Fimea. The Coordinating Ethics Committee of the Hospital District of Helsinki and Uusimaa (HUS) approved the FinnGen study protocol Nr HUS/990/2017.

The FinnGen study is approved by Finnish Institute for Health and Welfare (permit numbers: THL/2031/6.02.00/2017, THL/1101/5.05.00/2017, THL/341/6.02.00/2018, THL/2222/6.02.00/2018, THL/283/6.02.00/2019, THL/1721/5.05.00/2019, THL/1524/5.05.00/2020, and THL/2364/14.02/2020), Digital and population data service agency (permit numbers: VRK43431/2017-3, VRK/6909/2018-3, VRK/4415/2019-3), the Social Insurance Institution (permit numbers: KELA 58/522/2017, KELA 131/522/2018, KELA 70/522/2019, KELA 98/522/2019, KELA 138/522/2019, KELA 2/522/2020, KELA 16/522/2020 and Statistics Finland (permit numbers: TK-53-1041-17 and TK-53-90-20).

The Biobank Access Decisions for FinnGen samples and data utilized in FinnGen Data Freeze 6 include: THL Biobank BB2017_55, BB2017_111, BB2018_19, BB_2018_34, BB_2018_67, BB2018_71, BB2019_7, BB2019_8, BB2019_26, BB2020_1, Finnish Red Cross Blood Service Biobank 7.12.2017, Helsinki Biobank HUS/359/2017, Auria Biobank AB17-5154, Biobank Borealis of Northern Finland_2017_1013, Biobank of Eastern Finland 1186/2018, Finnish Clinical Biobank Tampere MH0004, Central Finland Biobank 1-2017, and Terveystalo Biobank STB 2018001.
